# The Selah trial: A preference-based partially randomized waitlist control study of three stress management interventions

**DOI:** 10.1101/2023.01.24.23284965

**Authors:** Rae Jean Proeschold-Bell, David E. Eagle, Logan C. Tice, Alyssa Platt, Jia Yao, Jessie S. Larkins, Eunsoo Timothy Kim, Joshua A. Rash

**Author notes:** **CORRESPONDING AUTHOR** Rae Jean Proeschold-Bell, PhD, Duke Global Health Institute, Duke Center for Health Policy & Inequalities Research, 310 Trent Drive, Room 310, Durham, NC 27705.

## Abstract

**Objective:** Chronic stress can undermine psychological and physiological health. We sought to evaluate three stress management interventions among clergy, accounting for intervention preferences.

**Methods:** United Methodist clergy in North Carolina enrolled in a partially randomized, preference-based waitlist control trial. The interventions were: mindfulness-based stress reduction (MBSR), Daily Examen prayer practice, and Stress Proofing (stress inoculation plus breathing skills). The intervention period spanned 12 weeks with a 12-week follow-up. Daily text message data were collected to assess practice across the 24 weeks. Co-primary outcomes were symptoms of stress using the Calgary Symptoms of Stress Inventory and 48-hour ambulatory heart rate variability (HRV) at 12-weeks post-intervention compared to waitlist control. Survey data were collected at 0, 12 and 24 weeks, with HRV collected at 0 and 12 weeks.

**Results:** 255 participants (mean age=54 years old; 91% white; 48% female) were randomized and initiated an intervention (n=184) or waitlist control (n=71). Compared to waitlist control, lower stress symptoms were found for MBSR participants [Mean Difference (MD)=-0.30, 95% CI:-0.41,-0.20; *p*<.001] and Stress Proofing (MD=-0.27, 95% CI:-0.40,-0.14; *p*<.001) at 12 weeks, and Daily Examen participants not until 24 weeks (MD=-0.24, 95% CI:-0.41,-0.08). Only MBSR participants demonstrated improvement in HRV at 12 weeks (MD=+3.32 millisecond; 95% CI:0.21,6.44; *p*=.036).

**Conclusions:** MBSR demonstrated robust improvement in self-reported and objective physical correlates of stress whereas Stress Proofing and Daily Examen resulted in improvements in self-reported correlates of stress only. These brief practices were sustainable and beneficial for an occupational sample during the COVID pandemic.

**Registration:** ClinicalTrials.gov identifier: NCT04625777 (https://clinicaltrials.gov/ct2/show/NCT04625777)

## 1 INTRODUCTION

Stress is a complex phenomenon occurring when the demands of a situation exceed the resources that one has to cope effectively [1]. Stress involves a physiological component (i.e., bodily changes) and a psychological component (e.g., perception of circumstances in life) [2]. When not managed well, stress can contribute to physiological problems (e.g., coronary artery disease, stroke, myocardial infarct) [3], [4] and psychological concerns (e.g., major depression) [5].

Mainline Protestant clergy exhibit average-to-elevated prevalence of chronic diseases [6,7] and symptoms of depression and anxiety [8, 9, 10, 11]. These physical and mental health concerns among clergy may relate to exposure to chronic stressors from an occupation that is emotionally demanding with leadership responsibilities, public criticism, and few breaks.

Stress management practices are needed that are acceptable and feasible for professionals engaged in challenging work. There is a diverse array of interventions developed for stress management, such as various kinds of meditation, diaphragmatic breathing, biofeedback, stress inoculation treatment, and cognitive behavioral therapy [12]. Of the available interventions, mindfulness-based stress reduction (MBSR) is well-established with demonstrated efficacy at improving symptoms of stress, anxiety, and negative emotions among diverse populations [13, 14, 15]. Unfortunately, poor engagement in mindfulness-based interventions can be a barrier to obtaining beneficial effects, and dropout rates can exceed 25% [16]. Moreover, individuals who engage in mindfulness-based interventions practice an average of 30 minutes per day on six days per week which represents 64% of the assigned daily practice duration [17]. This is problematic given that weak associations have been observed between self-reported home practice and intervention outcomes among 28 studies (*r*=.26, 95 %CI:.19-.34) [18]. Stress inoculation treatment has demonstrated efficacy for anxiety [19] and depression [20], although research on adherence to treatment over time is limited. Certain aspects of stress inoculation therapy, such as briefly engaging in stressful activities to gain confidence in one’s ability to endure stressors, draw on cognitive behavioral therapy, which reliably results in improvement of stress symptoms [21], [22].

Rather than adopt a one-size-fits-all approach, diverse stress management approaches may be needed to best meet the preferences of heterogeneous populations. Innovations in stress management practices may take several forms. One possibility is looking at the pathways (e.g., attention focused on the moment, non-judgmental thinking) that lead to stress symptom improvement for MBSR [23] and identify practices, such as prayer practices, that theoretically share those pathways. Another possibility is to take practices with an evidence base, such as stress inoculation training [24, 25] and package them in novel ways to potentially enhance uptake.

To assess stress management outcomes, stress can be thought of as a latent construct that varies temporally and requires assessment of multi-level indicators to adequately capture [26]. Stress can be measured as an exposure or a response, and can be approximated through behavioral symptoms or cognitive (e.g., appraisals), affective (e.g., mood and motivation), or physiological (e.g., operation of the autonomic nervous system). In this study, we sought to capture stress as a response through the use of measures that capture physical and psychological symptoms associated with chronic stress (e.g., downstream symptoms of physiological and affective processes), and a biological marker associated with stress [i.e., heart rate variability (HRV)].

HRV reflects variations in heart rate that index the capacity of the parasympathetic nervous system to alter heart rate in order to effectively meet the demands of a stressful event [27]. HRV has been shown to reliably covary with stress during stress-inducing procedures [28], and lower levels of HRV are reliably associated with depressed mood [29, 30], anxiety disorders [31], and reports of heightened occupational distress [32]. Further, HRV is a strong indicator of morbidity and risk of mortality in longitudinal studies [33, 34].

Specific to mindfulness-based interventions, a meta-analysis concluded that evidence for HRV improvements from the interventions was equivocal [35]. Of interest, only three studies that were included in this meta-analysis evaluated long-term (i.e., 24-hour) recordings of HRV, which may represent a better indicator of chronic stress.

This study aimed to evaluate the effects of three stress management interventions (i.e., MBSR; the Daily Examen prayer practice; and a set of stress inoculation skills entitled Stress Proofing) shown to be acceptable and feasible in a pilot study among clergy [removed for blinded review]. Outcomes of interest included symptoms of stress and HRV (co-primary outcomes), anxiety symptoms (secondary outcome), and depressive symptoms (exploratory outcome). An additional aim was to determine the effect of participant intervention preference on outcomes. We hypothesized that participants in each of the three active intervention conditions would experience improvements in each outcome compared to the waitlist control condition, and that participants with specific intervention preferences would experience larger improvements.

## 2 METHODS

### 2.1 Study design

We conducted a partially-randomized preference trial with a waitlist-control. Preference-based trials, a type of pragmatic clinical trial design, recognize that individuals have treatment preferences that are likely to affect outcomes due to expectancy effects or degree of engagement, which are particularly important in behavioral interventions [36]. In the partially randomized design, study participants are allowed to choose an intervention if they have a specific preference, whereas those without specific preferences are randomized to receive a particular intervention. Following initial enrollment in the partially-randomized preference trial, enrollment was re-opened after the start of the COVID-19 pandemic to an “observational” cohort whose participants were allowed to choose an intervention to participate in without a randomization structure (see study design details in Supplemental Methods Figure A1). The study protocol was registered under ClinicalTrials.gov identifier NCT04625777, and published online [37]. All procedures were approved by the Duke University Institutional Review Board and all participants gave informed consent.

### 2.2 Participants

Our study population was approximately 1,600 active United Methodist Church (UMC) clergy in North Carolina, USA, in 2019-2021. Most of these clergy served as congregational leaders, although some worked in non-profit and denominational settings, in rural, suburban, and urban environments. All clergy with a current appointment in one of the two North Carolina Annual Conferences were eligible if they were 18 years of age or older and willing to commit to completing their assigned stress management intervention. To enhance ecological validity, there were no stress-or health-related inclusion criteria.

### 2.3 Study procedures

#### 2.3.1 Recruitment and enrollment: trial cohort

We invited eligible clergy from November 2019 to January 2020 via mail and email addresses provided by the UMC conferences and announcements at in-person gatherings. We directed interested participants to information about the interventions and study details on a website, where they could begin the enrollment process prior to February 2020 by: 1) completing the Treatment Acceptability and Preferences Scale [38] for each intervention; 2) expressing their preferences among the three active interventions; and 3) providing consent for all study and intervention aspects. All of the randomized assignments were performed in February 2020, after all participants were enrolled. Participants assigned to immediate-intervention chose their desired intervention workshop dates from a list of available dates. The Selah trial cohort consists of participants who enrolled prior to March 1, 2020 and provided data while participating in an immediate intervention or the waitlist condition.

#### 2.3.2 Recruitment and enrollment: observational cohort

We re-opened enrollment in March 2020, anticipating that interest in stress management may increase with the start of the COVID-19 pandemic. Participants who enrolled after February 28, 2020 selected any intervention workshop and dates from a list; they were not randomized and represent an observational cohort that received their chosen intervention. The Selah observational cohort consists of these participants enrolled after March 1, 2020. We assigned participants who enrolled before March 1, 2020, to the waitlist condition, and after finishing the waitlist condition, they participated in an intervention and provided post-waitlist intervention data.

#### 2.3.3 Randomization

For the trial cohort, we asked participants whether they preferred any of the three interventions during the enrollment process. We randomized participants who stated no preference to be able to immediately receive one of the three active interventions or to the waitlist using an allocation ratio of 1:1:1:1. We randomized participants who preferred one intervention to immediately receive their preferred active intervention or to a waitlist with a 3:1 intervention vs waitlist ratio for MBSR and Stress Proofing, and a 5:4 intervention vs waitlist ratio for the Daily Examen (for all preference scenarios, see Supplemental Methods Table A1). The analysis statistician wrote code to generate the random allocation sequence in Stata version 16 [39]. Two staff members were responsible for accessing randomization results and informing participants of intervention allocation.

#### 2.3.4 Blinding

One staff member who was not the analysis statistician executed the randomization codes so that the analysis statistician could remain blinded to intervention allocation until data were collected and analysis decisions were finalized. Staff cleaning heart rate variability data were blind to intervention assignment. All others, including participants and intervention workshop instructors, were aware of group assignments. For details on methods for blinding the statistician during analysis, see Supplemental Appendix C.

#### 2.3.5 Intervention and data collection implementation

Implementation of the interventions and data collection for the trial and observational cohorts occurred during April 2020-October 2021. The three active interventions were designed as multi-session workshops and are described in detail in the protocol manuscript [cite masked for blind review].

#### 2.3.6 Study setting

As a pandemic-related modification to the trial, workshops were conducted synchronously online in groups of 5-25 participants rather than in-person. Adaptations to study design due to COVID-19 have been described in detail previously (cite masked for blind review). Participants provided data from their homes or personal settings.

### 2.4 Interventions

#### 2.4.1 Mindfulness-based Stress Reduction (MBSR)

MBSR teaches a set of meditation activities, along with attention to attitudes. We contracted with certified instructors from [masked for blind review] who used a criterion-standard mindfulness-based intervention based on Jon Kabat-Zinn’s model [40, 41]. In a synchronous web-based videoconference platform course, an instructor taught eight weekly 90-minute, synchronous, web-based sessions on awareness of breath, body scans, walking meditation, “choiceless” open awareness, Loving Kindness Meditation, yoga, and bringing awareness to the present moment. The course included study participants only and provided meditation instruction, periods of guided practice, and group discussion. Participants were asked to practice MBSR for 45 minutes per day for six months following their first of the eight sessions. After the eight sessions, participants were offered a four-hour online “Day of Mindfulness” which included participants and community members not enrolled in the study. Each series of eight sessions was taught by one of four certified instructors, who followed the same materials and order.

#### 2.4.2 The Daily Examen Prayer Practice

The Daily Examen is a Jesuit reflective prayer practice developed by Ignatius of Loyola and widely practiced by Christians from many traditions. For this study we used a modern adaptation of the Daily Examen [42].

The Daily Examen consists of a five-step prayer: 1) Become aware of God’s presence; 2) review the events of the past 24 hours, recalling 2-3 things for which you are grateful; 3) review the events of the past 24 hours, guided by the Holy Spirit, noticing where you experienced God’s presence; review what stands out and pay attention to what emotions arise. With the guidance of the Holy Spirit, pray through these emotions, noticing which are drawing you closer to God or pulling you away from God; and 5) look forward to the next 24 hours. What is one thing you should do? Where do you need God’s assistance?

With some similarities to mindfulness-based practices, these five steps help participants attend to the present by reflecting on positive emotions, moving past negative emotions, and aligning their actions with their perception of God’s wishes, and doing so with decreased judgment of their thoughts and feelings. The intervention consisted of three 90-minute, synchronous, web-based sessions on sequential days and involved practicing the Daily Examen, didactic content on the practice and practicalities of developing a daily prayer practice, and small group discussion. The sessions were each taught by two instructors. We asked participants to commit to practicing the Daily Examen daily for 10-15 minutes over 6 months following their workshop. Two and six weeks following their workshop, participants had the option to meet with their instructors in an online small group up to twice to address issues arising from their practice.

#### 2.4.3 Stress Proofing inoculation combination

Stress Proofing is a set of stress reduction skills with aspects of stress inoculation training [25, 43] selected and packaged by the NC Systema organization founder into four weekly 90-minute, synchronous, web-based sessions. The four-session series began with education on the stress response and awareness of one’s own stress response. The training diverged from traditional stress inoculation training and focused on physical activities to undo the stress response, including walking while diaphragmatic breathing, triangle and square breathing, tension control, stretching, and massage. The instructor discussed stress inoculation training and encouraged participants to embrace physical discomfort to learn to tolerate discomfort in the future [26]. The session content recommended a variety of beneficial lifestyle practices, including prioritizing nutrition and sleep and disengaging from technological devices an hour before sleep. We asked participants to practice diaphragmatic breathing and engage in any activity taught during Stress Proofing daily for six months. Workshops were taught by the founder; the founder had no involvement in the design, conduct, analysis, or reporting of the trial.

#### 2.4.4 Waitlist condition

Waitlist participants waited at least six months to participate in interventions, and during this time we asked them to complete surveys at 0-, 12-, and 24-weeks. We invited participants without disqualifying health conditions to additionally provide a 48-hour continuous ambulatory heart rate recording coinciding with their 0-and 12-week surveys. After completing the waiting period, we informed participants that they could update their intervention preference and receive an intervention while providing survey and HRV data.

### 2.5 Measures

#### 2.5.1 Co-primary outcomes

##### 2.5.1.1 Stress symptoms

Stress symptoms were measured using the subscales of anger, muscle tension, cardiopulmonary arousal, neurological/gastroenterological, and cognitive disorganization (total 41 items) of the Calgary-Symptoms of Stress Inventory (C-SOSI), a reliable and valid measure guided by mindfulness-based theory in its development [44, 45]. Participants were asked to indicate on a scale of 0 to 4 how often they experienced each symptom when presented with a stressor (0 = Never, 4 = Frequently). We used continuous mean scores of all the items (range 0-4), with higher mean scores indicating worse symptoms.

##### 2.5.1.2 Heart rate variability (HRV)

Ambulatory HRV was measured across a 48-hour period. Two individual-level cosine function parameters were estimated by Ordinary Least Squares regression to quantify the circadian variability parameters: 1) Midline Estimating Statistic Of Rhythm (MESOR), defined as the rhythm adjusted 24-hour mean, and 2) amplitude, defined as the distance between MESOR and the maximum of the cosine curve (i.e. half the extent of rhythmic change in a cycle). HRV procedures, including exclusion criteria, are detailed in the Supplemental Methods.

##### 2.5.2 Secondary and exploratory outcomes

The secondary outcome, symptoms of anxiety, was measured using the seven-item Generalized Anxiety Disorder-7 (GAD-7) scale (sum scores ranged from 0-21, with scores of ≥8 screening positive for elevated anxiety symptoms) [46, 47]. The exploratory outcome of depressive symptoms was measured using the eight-item Patient Health Questionnaire-8 (PHQ-8; sum scores ranged from 0-24, with scores of ≥10 screening positive for elevated depressive symptoms [48]. Psychometric properties of the GAD-7 and the PHQ-8 are well-documented [49].

#### 2.5.3 Demographic, intervention preference, and clinical measures

Sociodemographic and clinical measures were obtained by self-administered surveys during the baseline, 12-, and 24-week time periods. The clinically relevant constructs included physical activity, body mass index (BMI), caffeine and alcohol intake, and overall life stress level at enrollment. Preference measures were collected by self-administered surveys; the Treatment Acceptability and Preferences Scale [50] was administered during the enrollment period and preference for online vs in-person intervention was administered at baseline. Complete details are provided in the protocol paper [removed for blinded review] and Supplemental Methods.

#### 2.5.4 Engagement measures

##### 2.5.4.1 Daily practice reports via text

We sent participants a daily text message for 24 weeks during the active intervention period. We asked MBSR participants for the number of minutes practiced during the prior day. We asked Daily Examen participants whether or not they had practiced during the prior day. We asked Stress Proofing participants if they had conducted 0, 1, or 2 “resets” (i.e., Stress Proofing practices) during the prior day.

### 2.6 Data collection procedures

#### 2.6.1 Intervention participants

Data collection procedures were the same for trial and observational cohort participants. We collected survey data, solicited by email and administered in REDCap database software 12.4.28 [51], at intervention start (0 weeks), 12 weeks, and 24 weeks. We collected heart rate variability data at 0 and 12 weeks. We mailed participants Bittium eMotion Faros 180 ambulatory heart rate recording devices and invited them to a synchronous learning session held via webplatform and additionally directed them to an online instructional video. After collecting their 48-hour sample, participants returned their devices by mail. Study staff downloaded the data and cleaned and processed it using Kubios Premium software 3.4.1 [52]. To send and receive text messages, the study used a software engine designed by Duke software engineers, called Prompt. Prompt receives self-monitoring data that participants send via text message. Prompt interfaces with Twilio software [53] to process data for the engine. Self-monitoring data were automatically analyzed each week according to embedded algorithms. Baseline data collection occurred after randomization among trial participants, thus it was possible for dropout to have occurred prior to baseline data collection.

#### 2.6.2 Waitlist participants

We initiated data collection from waitlist participants in groups of 20 starting in June 2020, July 2020, September 2020, and February 2021 to span the range of data collection from active intervention participants. The spacing of data collection during the waiting period matched that of intervention participants: surveys at 0, 12, and 24 weeks, and HRV data at 0 and 12 weeks. We asked waitlist participants who proceeded to start an intervention following the waiting period to provide data again during their intervention period following this same schedule. We included the post-waitlist intervention data in sensitivity analyses.

#### 2.6.3 Incentives

Data collection was incentivized. We compensated participants $25 for each occasion of 48-hour ambulatory HRV data submitted, $20 each for 0-and 12-week surveys, and $25 for 24-week surveys.

#### 2.6.4 Data availability

The datasets generated during the current study are not publicly available but we will make de-identified data available for reasonable requests compliant with ethical approvals from the sending and receiving hosts’ institutional ethics review boards.

### 2.7 Sample size

Based on results from our non-randomized pilot study (conducted prior to the full trial), we expected an average baseline C-SOSI score of 0.92 (SD=0.46) across all interventions, with 12-week follow-up scores of 0.7 (SD=0.58) for MBSR, 0.55 (SD=0.36) for Stress Proofing, and 0.51 (SD=0.38) for the Daily Examen. A per-arm sample size of 40 for Daily Examen, 47 for Stress Proofing, and 195 for MBSR (which had a larger standard deviation, resulting in a larger sample size) would have 80% power to detect a similar between-arm differences in means as were observed in the pilot study, using a two-sample t-test with unequal variance and factoring in an expected 20% loss to follow-up and design effect of 1.3 (corresponding to an ICC of 0.027 and an average cluster size of 12) due to clustering caused by group delivery of the intervention. For HRV, based on previous literature recommending a standardized mean difference of 0.50 for a medium effect size [54], a per-arm sample size of 140 would have 80% power to detect a medium effect size for a two-sample t-test, assuming a similar follow-up rate and design effect as with C-SOSI. All sample size calculations assumed an alpha of 0.0167 based on a Bonferroni correction on 3 hypotheses (for 3 interventions) and were conducted using PASS 2021 software. Additional details on the power analysis inputs and methods can be found in the Supplementary Methods Appendix.

### 2.8 Statistical analysis

The Selah study team administers a biennial panel survey of all UMC clergy in North Carolina [removed for blinded review]. The 2019 wave of the panel survey obtained a 73% response rate and provided a large sample of the general population of clergy that were eligible to participate in Selah. We compared descriptive statistics between the Selah trial phase and the 2019 panel survey to determine characteristics associated with self-selection of clergy into the Selah study and to inform generalizability of results.

Use of the partially randomized preference design during the trial period meant that by design the analytic data would be a mix of randomized data (for trial participants that had no preference) and observational data (for trial participants that had a preference and were allowed to select their intervention), which made it likely that treatment arms would be imbalanced on baseline characteristics in ways that may have been associated with study outcomes in an unadjusted analysis. In addition, randomization was performed prior to baseline data collection, with substantial dropout before data collection commenced. A propensity score covariate adjustment method [55] was selected using covariate balancing propensity scores [56]; details are provided in the Supplemental Methods. All analyses use an as-treated approach to calculate treatment effects.

Main outcome models used linear mixed effects models with random intercepts at the level of the individual to account for repeated measurements within individuals. Random slopes on a binary treatment indicator for the group assignment were used to produce a random intercept for each workshop and a separate intercept for the un-clustered control arm in order to account for partial clustering due to group-administered treatment [57]. We calculated an interclass correlation coefficient (ICC) for each outcome model to quantify the level of clustering due to group treatment. We included a cubic functional form for calendar time to protect against time confounding (details on justification and methods included in Supplemental Methods).

Because propensity score adjustment was used to balance baseline levels of the outcomes (in addition to other prognostic characteristics), we chose to model treatment effects longitudinally using a constrained longitudinal data analysis modeling (cLDA) approach, which models baseline as an outcome and assumes baseline levels of the outcome are equal across arms [58]. Due to known variation in timing of the 12-and 24-week surveys, time was modeled continuously in weeks from baseline using linear splines with knots at 12 and 24 weeks for C-SOSI, GAD-7, and PHQ-8 outcomes and one spline at 12 weeks for HRV outcomes. Treatment effects were between arm differences in outcomes at 12 weeks (primary) and 24 weeks (secondary). We used robust sandwich standard errors to account for the fact that propensity scores were estimated.

We extracted and compiled the text data to calculate the proportion of participants engaging in their assigned practice on each day throughout the 24 weeks. Analysis of practice data was purely descriptive with no hypothesis testing.

#### 2.8.1 Subgroup Analysis

Subgroup analysis was performed to ascertain whether treatment effects were different for participants who received an intervention that they uniquely preferred versus those that had tied or no preference or who received an intervention other than the one for which they expressed a unique preference.

Treatment effect estimates by preference status were calculated using binary interaction terms with treatment and time terms with calculated linear combinations for treatment estimate by preference status. These subgroup analyses are considered strictly exploratory, thus analytic focus should be on effect estimates, confidence intervals, and magnitude of interaction terms rather *p*-values. Any findings of heterogeneity or lack thereof would need confirmation via a future prospective study in order to make definitive conclusions.

#### 2.8.2 Sensitivity analysis incorporating post-trial period data

Data collected from trial participants who provided post-waitlist data while receiving an intervention or from observational cohort participants who enrolled in the study after March 1, 2020 were considered fully observational and separate from the trial and thus excluded from the main analysis. However, a sensitivity analysis was performed pooling these data with the trial data to ascertain whether results remained when all available data were used.

#### 2.8.3 Missing Data

Missing data were present both in baseline covariates used to generate propensity scores and 12-and 24-week outcome data due to study dropout. Therefore, sensitivity analyses were performed using multiple imputation with chained equations (MICE) [59] (see Supplement) to assess the extent to which missing data and study dropout may have affected the magnitude and direction of treatment effect estimates. Propensity scores were calculated separately for each of the imputation datasets and estimates combined using Rubin’s rules [60].

All statistical tests used an alpha of 0.05. Because we were interested in examining the effectiveness of each intervention separately and there were two correlated primary outcomes of interest (C-SOSI and HRV MESOR), *p*-values were adjusted for two tests separately for each intervention [61] based on trial data using the Benjamini-Hochberg procedure [62]. Original and corrected *p*-values are presented only for primary outcomes.

## 3 RESULTS

### 3.1 Study flow

As shown in Supplemental Figure B1, 1,642 eligible UMC clergy were invited to participate; prior to March 1, 2020, 390 consented. Assignment to interventions was preference-based with 310 (79.5%) indicating a preference for one single intervention or ambivalence between two interventions, and 80 (20.5%) indicating no preference among the three interventions. Of the 310 participants with singular preference or tied between two interventions, 207 were assigned to an immediate intervention (with 144 going forward to participate in interventions), while 103 were assigned to the waitlist (with 62 providing baseline data). Among the 80 without any preference, 60 were assigned to immediate intervention (of whom 40 went on to participate and provide baseline data) and 20 were assigned to the waitlist (with 9 providing baseline data). Of the 255 participants providing survey data in the trial period, 157 also provided HRV data; 8% (n=20) were excluded from HRV data collection and 31% (n=78) had missing data. See Supplement Table A2 for exclusion reasons and counts.

Among the 255 participants, 174 stated a unique intervention preference at study registration and received the same intervention during the trial period. Reasons for not receiving one’s initial preferred intervention were participant-driven: their preference changed during the time between enrollment and intervention start, possibly due to new circumstances from the pandemic, our switch from in-person to online-only delivery, or logistical reasons such as specific intervention dates and times. As shown in Supplement Figure B1, 47 participants randomized to waitlist participated in post-waitlist interventions and were included in sensitivity analyses. An additional 63 individuals consented to participate in the interventions after March 1, 2020 (see Supplemental Figure B2), of whom 50 participated and were included in sensitivity analyses.

### 3.2 Sample Characteristics

Table 1 reports comparisons of baseline characteristics between participants assigned to the immediate interventions versus waitlist. Participants were evenly split between females (47.5%) and males (52.5%), with a mean age of 53.9 (SD=11.2) years. Participants were predominantly white (90.6%) or Black (5.9%), married or cohabitating (89.4%), and serving a church (82.4%). Supplemental Tables B2 and B3 report baseline characteristics of the subsample of participants that provided HRV data and pooled trial and observational participants.

**Table 1.**
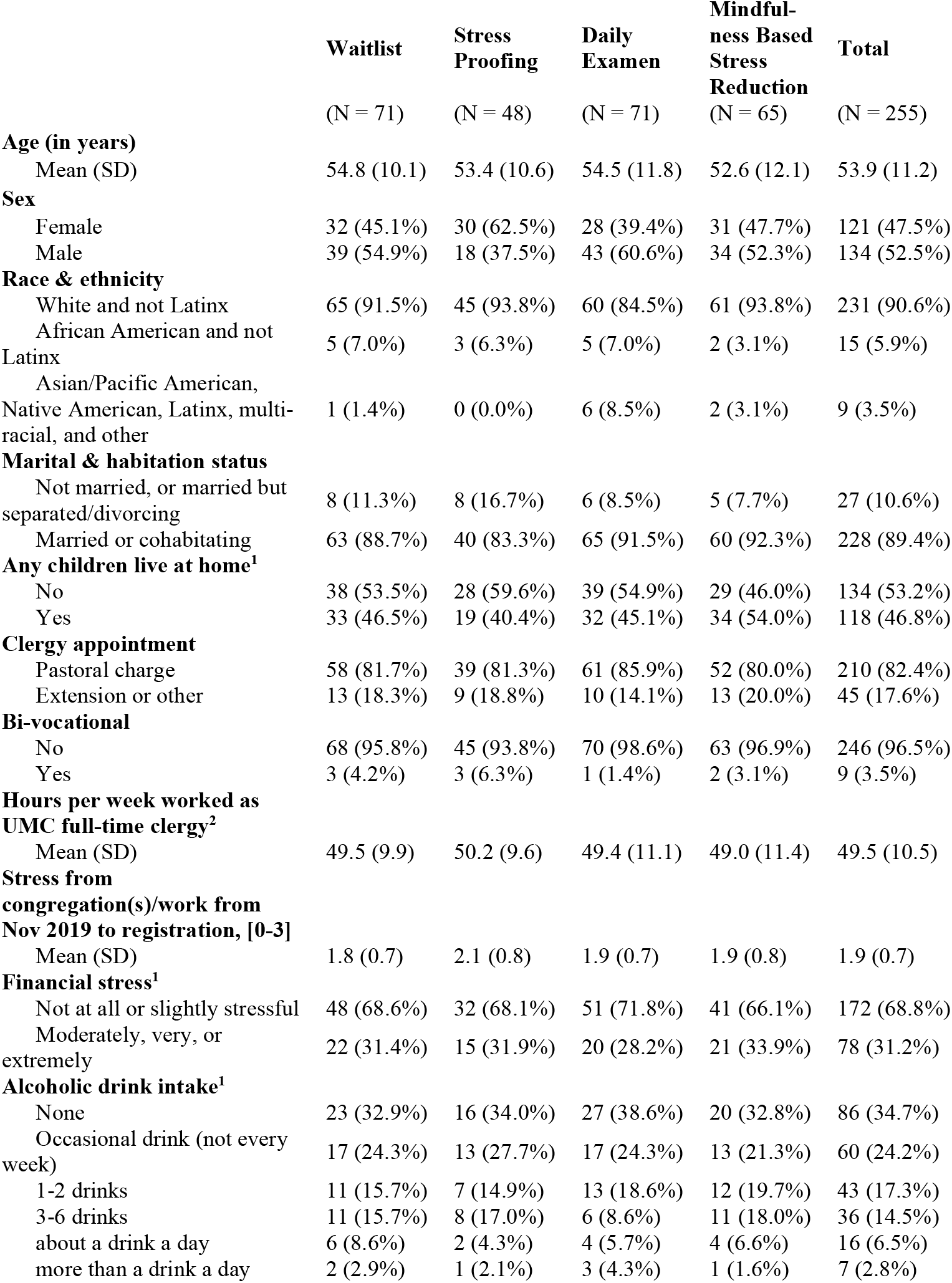
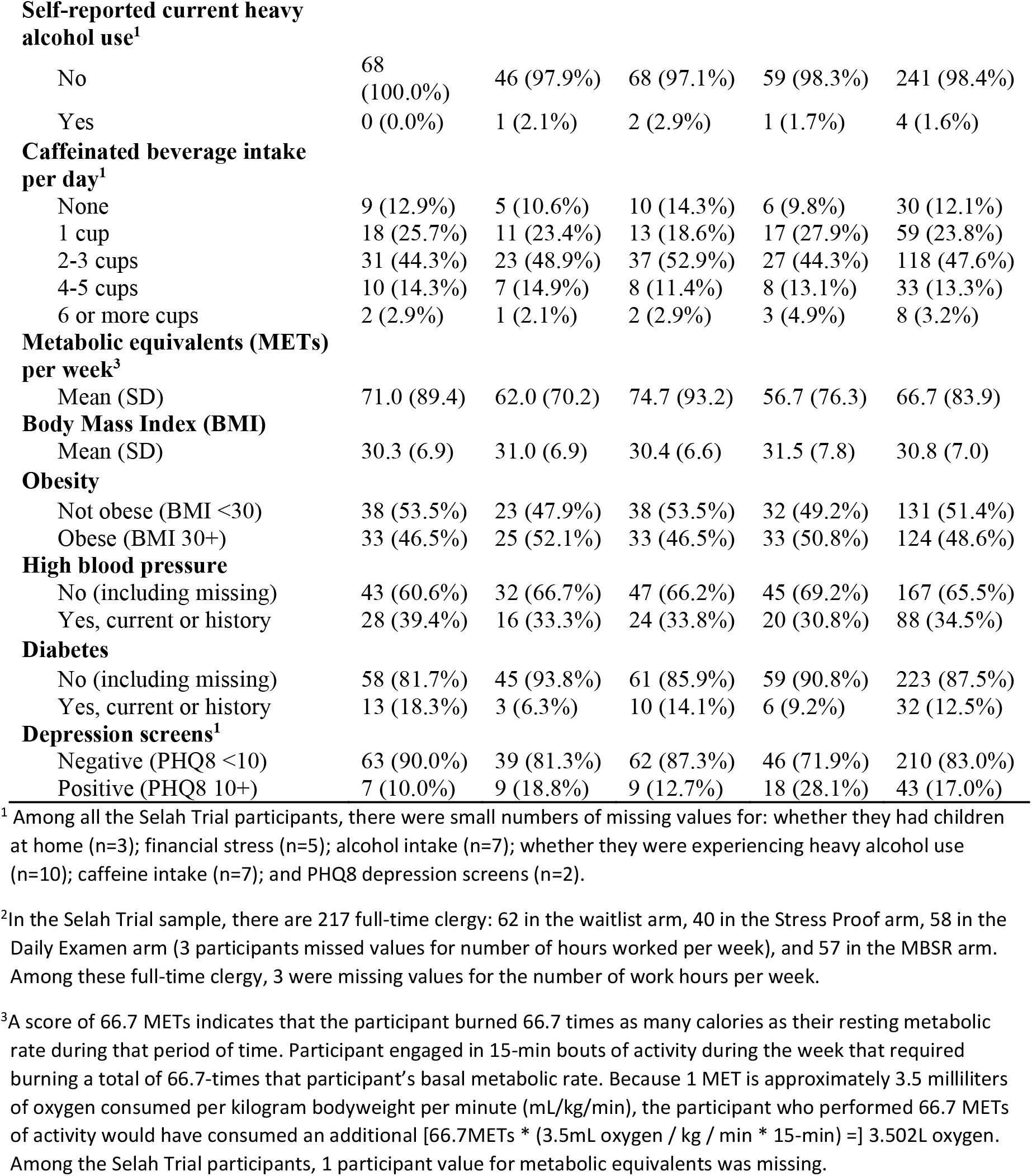
Baseline characteristics of active intervention and waitlist study arms for trial periods participants

|  | **Waitlist** | **Stress**<br>**Proofing** | **Daily**<br>**Examen** | **Mindful-**<br>**ness Based**<br>**Stress**<br>**Reduction** | **Total** |
| --- | --- | --- | --- | --- | --- |
|  | (N = 71) | (N = 48) | (N = 71) | (N = 65) | (N = 255) |
| **Age (in years)** |  |  |  |  |  |
| Mean (SD) | 54.8 (10.1) | 53.4 (10.6) | 54.5 (11.8) | 52.6 (12.1) | 53.9 (11.2) |
| **Sex** |  |  |  |  |  |
| Female | 32 (45.1%) | 30 (62.5%) | 28 (39.4%) | 31 (47.7%) | 121 (47.5%) |
| Male | 39 (54.9%) | 18 (37.5%) | 43 (60.6%) | 34 (52.3%) | 134 (52.5%) |
| **Race & ethnicity** |  |  |  |  |  |
| White and not Latinx | 65 (91.5%) | 45 (93.8%) | 60 (84.5%) | 61 (93.8%) | 231 (90.6%) |
| African American and not Latinx | 5 (7.0%) | 3 (6.3%) | 5 (7.0%) | 2 (3.1%) | 15 (5.9%) |
| Asian/Pacific American, Native American, Latinx, multi-racial, and other | 1 (1.4%) | 0 (0.0%) | 6 (8.5%) | 2 (3.1%) | 9 (3.5%) |
| **Marital & habitation status** |  |  |  |  |  |
| Not married, or married but separated/divorcing | 8 (11.3%) | 8 (16.7%) | 6 (8.5%) | 5 (7.7%) | 27 (10.6%) |
| Married or cohabitating | 63 (88.7%) | 40 (83.3%) | 65 (91.5%) | 60 (92.3%) | 228 (89.4%) |
| **Any children live at home<sup>1</sup>** |  |  |  |  |  |
| No | 38 (53.5%) | 28 (59.6%) | 39 (54.9%) | 29 (46.0%) | 134 (53.2%) |
| Yes | 33 (46.5%) | 19 (40.4%) | 32 (45.1%) | 34 (54.0%) | 118 (46.8%) |
| **Clergy appointment** |  |  |  |  |  |
| Pastoral charge | 58 (81.7%) | 39 (81.3%) | 61 (85.9%) | 52 (80.0%) | 210 (82.4%) |
| Extension or other | 13 (18.3%) | 9 (18.8%) | 10 (14.1%) | 13 (20.0%) | 45 (17.6%) |
| **Bi-vocational** |  |  |  |  |  |
| No | 68 (95.8%) | 45 (93.8%) | 70 (98.6%) | 63 (96.9%) | 246 (96.5%) |
| Yes | 3 (4.2%) | 3 (6.3%) | 1 (1.4%) | 2 (3.1%) | 9 (3.5%) |
| **Hours per week worked as UMC full-time clergy<sup>2</sup>** |  |  |  |  |  |
| Mean (SD) | 49.5 (9.9) | 50.2 (9.6) | 49.4 (11.1) | 49.0 (11.4) | 49.5 (10.5) |
| **Stress from congregation(s)/work from Nov 2019 to registration, [0-3]** |  |  |  |  |  |
| Mean (SD) | 1.8 (0.7) | 2.1 (0.8) | 1.9 (0.7) | 1.9 (0.8) | 1.9 (0.7) |
| **Financial stress<sup>1</sup>** |  |  |  |  |  |
| Not at all or slightly stressful | 48 (68.6%) | 32 (68.1%) | 51 (71.8%) | 41 (66.1%) | 172 (68.8%) |
| Moderately, very, or extremely | 22 (31.4%) | 15 (31.9%) | 20 (28.2%) | 21 (33.9%) | 78 (31.2%) |
| **Alcoholic drink intake<sup>1</sup>** |  |  |  |  |  |
| None | 23 (32.9%) | 16 (34.0%) | 27 (38.6%) | 20 (32.8%) | 86 (34.7%) |
| Occasional drink (not every week) | 17 (24.3%) | 13 (27.7%) | 17 (24.3%) | 13 (21.3%) | 60 (24.2%) |
| 1-2 drinks | 11 (15.7%) | 7 (14.9%) | 13 (18.6%) | 12 (19.7%) | 43 (17.3%) |
| 3-6 drinks | 11 (15.7%) | 8 (17.0%) | 6 (8.6%) | 11 (18.0%) | 36 (14.5%) |
| about a drink a day | 6 (8.6%) | 2 (4.3%) | 4 (5.7%) | 4 (6.6%) | 16 (6.5%) |
| more than a drink a day | 2 (2.9%) | 1 (2.1%) | 3 (4.3%) | 1 (1.6%) | 7 (2.8%) |
| **Self-reported current heavy alcohol use<sup>1</sup>** |  |  |  |  |  |
| No | 68<br>(100.0%) | 46 (97.9%) | 68 (97.1%) | 59 (98.3%) | 241 (98.4%) |
| Yes | 0 (0.0%) | 1 (2.1%) | 2 (2.9%) | 1 (1.7%) | 4 (1.6%) |
| **Caffeinated beverage intake per day<sup>1</sup>** |  |  |  |  |  |
| None | 9 (12.9%) | 5 (10.6%) | 10 (14.3%) | 6 (9.8%) | 30 (12.1%) |
| 1 cup | 18 (25.7%) | 11 (23.4%) | 13 (18.6%) | 17 (27.9%) | 59 (23.8%) |
| 2-3 cups | 31 (44.3%) | 23 (48.9%) | 37 (52.9%) | 27 (44.3%) | 118 (47.6%) |
| 4-5 cups | 10 (14.3%) | 7 (14.9%) | 8 (11.4%) | 8 (13.1%) | 33 (13.3%) |
| 6 or more cups | 2 (2.9%) | 1 (2.1%) | 2 (2.9%) | 3 (4.9%) | 8 (3.2%) |
| **Metabolic equivalents (METs) per week<sup>3</sup>** |  |  |  |  |  |
| Mean (SD) | 71.0 (89.4) | 62.0 (70.2) | 74.7 (93.2) | 56.7 (76.3) | 66.7 (83.9) |
| **Body Mass Index (BMI)** |  |  |  |  |  |
| Mean (SD) | 30.3 (6.9) | 31.0 (6.9) | 30.4 (6.6) | 31.5 (7.8) | 30.8 (7.0) |
| **Obesity** |  |  |  |  |  |
| Not obese (BMI <30) | 38 (53.5%) | 23 (47.9%) | 38 (53.5%) | 32 (49.2%) | 131 (51.4%) |
| Obese (BMI 30+) | 33 (46.5%) | 25 (52.1%) | 33 (46.5%) | 33 (50.8%) | 124 (48.6%) |
| **High blood pressure** |  |  |  |  |  |
| No (including missing) | 43 (60.6%) | 32 (66.7%) | 47 (66.2%) | 45 (69.2%) | 167 (65.5%) |
| Yes, current or history | 28 (39.4%) | 16 (33.3%) | 24 (33.8%) | 20 (30.8%) | 88 (34.5%) |
| **Diabetes** |  |  |  |  |  |
| No (including missing) | 58 (81.7%) | 45 (93.8%) | 61 (85.9%) | 59 (90.8%) | 223 (87.5%) |
| Yes, current or history | 13 (18.3%) | 3 (6.3%) | 10 (14.1%) | 6 (9.2%) | 32 (12.5%) |
| **Depression screens<sup>1</sup>** |  |  |  |  |  |
| Negative (PHQ8 <10) | 63 (90.0%) | 39 (81.3%) | 62 (87.3%) | 46 (71.9%) | 210 (83.0%) |
| Positive (PHQ8 10+) | 7 (10.0%) | 9 (18.8%) | 9 (12.7%) | 18 (28.1%) | 43 (17.0%) |

<sup>1</sup> Among all the Selah Trial participants, there were small numbers of missing values for: whether they had children at home (n=3); financial stress (n=5); alcohol intake (n=7); whether they were experiencing heavy alcohol use (n=10); caffeine intake (n=7); and PHQ8 depression screens (n=2).
<sup>2</sup> In the Selah Trial sample, there are 217 full-time clergy: 62 in the waitlist arm, 40 in the Stress Proof arm, 58 in the Daily Examen arm (3 participants missed values for number of hours worked per week), and 57 in the MBSR arm. Among these full-time clergy, 3 were missing values for the number of work hours per week.
<sup>3</sup> A score of 66.7 METs indicates that the participant burned 66.7 times as many calories as their resting metabolic rate during that period of time. Participant engaged in 15-min bouts of activity during the week that required burning a total of 66.7-times that participant's basal metabolic rate. Because 1 MET is approximately 3.5 milliliters of oxygen consumed per kilogram bodyweight per minute (mL/kg/min), the participant who performed 66.7 METs of activity would have consumed an additional [66.7METs \* (3.5mL oxygen / kg / min \* 15-min) =] 3.502L oxygen. Among the Selah Trial participants, 1 participant value for metabolic equivalents was missing.

Supplemental Table B1 depicts comparisons of characteristics of Selah trial phase participants with those of the eligible population, using 2019 Panel Study data. Females were more likely to participate in the Selah study than males (47.5% in Selah vs. 33.7% in the panel study, *p*<.001). Those who were bi-vocational (*p*<.001), had body mass index (BMI)<30 (*p*=.043), and self-reported diabetes (*p*=.002) were less likely to participate in the Selah study, and those with elevated depressive symptoms were more likely to participate (*p=*.005).

### 3.3 Engagement

Participation in intervention sessions was high across all three active interventions (Supplemental Table B4). The median size of intervention groups was eight (IQR: 4,10) across a total of 21 groups. For Stress Proofing, 87.5% attended three out of four main sessions and 62.5% of participants attended four out of four main sessions, with more than half attending the optional follow-up session. For the Daily Examen, 95.8% of participants attended all three sessions, with more than half attending at least one optional follow-up session. For MBSR, the median participant attended seven of eight sessions.

The text message response rate for all interventions peaked in the first three weeks at an overall response rate of 88.0%, (Stress Proofing, 85.9%; Daily Examen, 90.2%; MBSR, 86.5%; Supplemental Figure B3). By 24 weeks, text response rates had fallen to 70.0%. Those reporting that they had practiced any MBSR declined slowly but steadily over the 24-week period, with the average reported minutes of practice per day (for those reporting any practice) across the 24 weeks being 28.4 (16.8) minutes.

### 3.4 Propensity scores

Supplemental Table A1 shows the variable specifications for propensity score models. Distributions of propensity scores indicated good overlap, with overdispersion at low propensity scores across all intervention types (Supplemental Figure B4). Comparisons of unweighted descriptive statistics between treatment conditions indicated relatively good a priori balance between participants in most sociodemographic characteristics. Those in Stress Proofing and MBSR exhibited higher baseline levels of stress, anxiety, and depressive symptoms than their counterparts in Daily Examen and the waitlist (Supplemental Table B5). After propensity score adjustment, differences between treatment arms were systematically reduced.

### 3.5 Primary Outcome Analyses

#### 3.5.1 Stress Symptoms (C-SOSI)

The baseline adjusted mean C-SOSI score across all arms was 1.01 (95% CI: 0.95, 1.06) (Table 2). Mean differences in C-SOSI scores between an active treatment arm and waitlist at 12 weeks were most pronounced for those participating in MBSR [Mean Difference (MD)=-0.30, 95% CI:-0.41,-0.20; *p*<.001] and Stress Proofing (MD= -0.27, 95% CI: -0.40, -0.14; *p*<.001), with less evidence of substantial differences for Daily Examen participants (MD= -0.08, 95% CI: -0.21, 0.05). By 24-weeks post-baseline, differences between active treatment arms and the waitlist control grew more substantial, with stronger evidence of differences between Daily Examen and waitlist participants (MD= -0.24, 95% CI: - 0.41, -0.08), in addition to larger differences between Stress Proofing and MBSR versus waitlist participants.

**Table 2.** Between arm, mixed effects regression estimated differences in outcomes between immediate intervention and waitlist by follow-up time point for trial period participants

|  | Sample size |  | Time point | Stress Proofing |  | Daily Examen |  | Mindfulness-Based Stress Reduction |  |
| --- | --- | --- | --- | --- | --- | --- | --- | --- | --- |
|  | Participants | Observations |  | Unadjusted | Adjusted | Unadjusted | Adjusted | Unadjusted | Adjusted |
| Survey outcomes |  |  |  |  |  |  |  |  |  |
| C-SOSI<br>(scale range 0-4) | 233 | 603 | Baseline | 1.00 | 1.01 | 1.00 | 1.01 | 1.00 | 1.01 |
|  |  |  | Mean<sup>1</sup> | [0.93, 1.07] | [0.95, 1.06] | [0.93, 1.07] | [0.95, 1.06] | [0.93, 1.07] | [0.95, 1.06] |
|  |  |  |  | -0.28 | -0.27 | -0.07 | -0.08 | -0.27 | -0.30 |
|  |  |  | 12 weeks | [-0.41, -0.16] | [-0.40, -0.14]<sup>2</sup> | [-0.21, 0.07] | [-0.21, 0.05]<sup>3</sup> | [-0.39, -0.16] | [-0.41, -0.20]<sup>4</sup> |
|  |  |  |  | -0.31 | -0.31 | -0.22 | -0.24 | -0.37 | -0.42 |
|  |  |  | 24 weeks | [-0.49, -0.13] | [-0.48, -0.14] | [-0.40, -0.05] | [-0.41, -0.08] | [-0.50, -0.23] | [-0.55, -0.28] |
| GAD-7<br>(scale range 0-21) | 233 | 603 | Baseline | 4.89 | 4.92 | 4.89 | 4.92 | 4.89 | 4.92 |
|  |  |  | Mean<sup>1</sup> | [4.37, 5.41] | [4.50, 5.34] | [4.37, 5.41] | [4.50, 5.34] | [4.37, 5.41] | [4.50, 5.34] |
|  |  |  |  | -1.22 | -1.29 | -0.47 | -0.51 | -1.62 | -1.85 |
|  |  |  | 12 weeks | [-2.18, -0.26] | [-2.27, -0.31] | [-1.37, 0.44] | [-1.42, 0.40] | [-2.43, -0.80] | [-2.66, -1.04] |
|  |  |  |  | -1.33 | -1.45 | -1.33 | -1.36 | -2.13 | -2.40 |
|  |  |  | 24 weeks | [-2.55, -0.10] | [-2.68, -0.23] | [-2.48, -0.17] | [-2.49, -0.24] | [-3.21, -1.05] | [-3.43, -1.36] |
| PHQ-8<br>(scale range 0-24) | 232 | 600 | Baseline | 5.43 | 5.51 | 5.43 | 5.51 | 5.43 | 5.51 |
|  |  |  | Mean<sup>1</sup> | [4.86, 6.01] | [5.11, 5.91] | [4.86, 6.01] | [5.11, 5.91] | [4.86, 6.01] | [5.11, 5.91] |
|  |  |  |  | -1.82 | -1.72 | -1.22 | -1.21 | -2.06 | -2.07 |
|  |  |  | 12 weeks | [-2.95, -0.69] | [-2.82, -0.63] | [-2.39, -0.05] | [-2.37, -0.05] | [-3.13, -0.98] | [-3.15, -1.00] |
|  |  |  |  | -1.85 | -1.60 | -1.52 | -1.48 | -2.46 | -2.46 |
|  |  |  | 24 weeks | [-2.96, -0.74] | [-3.00, -0.20] | [-2.87, -0.18] | [-2.88, -0.08] | [-3.62, -1.31] | [-3.69, -1.24] |
| HRV outcomes |  |  |  |  |  |  |  |  |  |
| MESOR<br>(unit = 1 millisecond) | 142 | 253 | Baseline | 24.6 | 24.6 | 24.6 | 24.6 | 24.6 | 24.6 |
|  |  |  | Mean<sup>1</sup> | [22.2, 27.1] | [22.2, 27.1] | [22.2, 27.1] | [22.2, 27.1] | [22.2, 27.1] | [22.2, 27.1] |
|  |  |  |  | -1.99 | -0.64 | 1.64 | 1.38 | 3.31 | 3.32 |
|  |  |  | 12 weeks | [-5.33, 1.35] | [-3.92, 2.65]<sup>5</sup> | [-2.81, 6.08] | [-2.75, 5.52]<sup>6</sup> | [0.01, 6.61] | [0.21, 6.44]<sup>7</sup> |
| Amplitude<br>(unit = 1 millisecond) | 142 | 253 | Baseline | 7.74 | 7.88 | 7.74 | 7.88 | 7.74 | 7.88 |
|  |  |  | Mean<sup>1</sup> | [6.34, 9.14] | [6.54, 9.22] | [6.34, 9.14] | [6.54, 9.22] | [6.34, 9.14] | [6.54, 9.22] |
|  |  |  |  | 0.84 | 1.33 | 1.21 | 1.08 | 1.86 | 1.94 |
|  |  |  | 12 weeks | [-1.13, 2.81] | [-0.68, 3.33] | [-0.80, 3.21] | [-0.68, 2.84] | [0.07, 3.65] | [0.17, 3.72] |

Depending on the survey outcome, 46 Stress Proofing participants, 63 Daily Examen participants, and 57-58 MBSR participants were compared to 66 waitlist control participants. For each of the HRV outcomes, 30 Stress Proofing participants, 36 Daily Examen participants, and 37 MBSR participants were compared to 39 waitlist control participants.
Abbrev: C-SOSI = Calgary Symptoms of Stress Inventory; GAD = Generalized Anxiety Disorder scale; PHQ = Patient Health Questionnaire; HRV = Heart rate variability; MESOR = Midline estimating statistic of rhythm
<sup>1</sup>Regression estimated mean rather than difference between arms. Means are constrained to be the same across all arms at baseline
<sup>2</sup> $p < .001$ before and after Benjamini-Hochberg correction
<sup>3</sup> $p = .222$ before correction; $p = .445$ after correction
<sup>4</sup> $p < .001$ before and after correction
<sup>5</sup> $p = .704$ before and after correction
<sup>6</sup> $p = .511$ before and after correction
<sup>7</sup> $p = .036$ before and after correction

#### 3.5.2 Heart Rate Variability (HRV)

Baseline adjusted mean MESOR across all arms was 24.6 ms (95% CI: 22.2, 27.1) and amplitude was 7.88 ms (95% CI: 6.54, 9.22, Table 2). Participants in MBSR had a modest 3.32 ms higher mean MESOR (95% CI: 0.21, 6.44; *p*=.036) and a 1.94 ms higher amplitude (95% CI: 0.17, 3.72) than similar participants in waitlist control at 12 weeks. There was no evidence of a difference in MESOR or amplitude for the other intervention arms vs the waitlist.

### 3.6 Secondary and Exploratory Outcome Analyses

#### 3.6.1 Anxiety (GAD-7)

The baseline adjusted mean GAD-7 score across all arms was 4.92 (95% CI: 4.50, 5.34, Table 2). Similar to the stress symptoms results, participants in Stress Proofing (MD= -1.29 points, 95% CI: -2.27, -0.26 at 12 weeks; MD= -1.45, 95% CI: -2.68, -0.23 at 24 weeks) and MBSR (MD= -1.85 points, 95% CI: -2.66, -1.04 at 12 weeks; MD= -2.40, 95% CI: -3.43, -1.36 at 24 weeks) had fewer symptoms of anxiety at 12 and 24 weeks than comparable participants in the waitlist control, with the most pronounced differences for MBSR. Differences between Daily Examen and waitlist participants were modest at 12 weeks (MD=-0.51, 95% CI: -1.42, 0.40) and became more pronounced at 24 weeks (MD=-1.36, 95% CI: -2.49, -0.24).

#### 3.6.2 Depression (PHQ-8)

The baseline adjusted mean PHQ-8 score across all arms was 5.51 (95% CI: 5.11, 5.91, Table 2). Similar to the results for anxiety symptoms, Stress Proofing and MBSR participants had fewer symptoms of depression at 12 weeks than comparable participants in the waitlist control (Stress Proofing: MD= -1.72, 95% CI: -2.82, -0.63; MBSR: MD= -2.07, 95% CI: -3.15, -1.00), with sustained differences between participants in each intervention vs waitlist controls at 24 weeks (Stress Proofing: MD= -1.60, 95% CI: - 3.00, -0.20; MBSR: MD= -2.46, 95% CI: -3.69, -1.24). Differences between Daily Examen and waitlist participants were observed at both 12 weeks (MD=-1.21, 95% CI: -2.37, -0.05) and 24 weeks (MD=-1.48, 95% CI: -2.88, -0.08).

### 3.7 Subgroup analyses

Participants who received a uniquely preferred intervention (n=174) largely resembled those who did not (n=81) in terms of baseline sociodemographic and clinical characteristics. Being white (p=0.01), having a lower BMI (p=0.001), and having higher levels of physical activity (p=0.015) were correlated with receiving an initially preferred intervention (Supplemental Table B11).

There was little evidence of heterogeneity in treatment effects for stress, anxiety, and depressive symptom outcomes at 12 weeks when estimates were stratified by receipt of an initially uniquely preferred intervention (Figure 1, Supplemental Figure B5). For Stress Proofing outcomes at 24 weeks, there was some evidence that those whose initial preference did not match their intervention group had a larger treatment effect (lower stress and depressive symptoms compared to waitlist) than those whose initial unique preference matched their intervention group (stress symptoms interaction effect = 0.20, 95% CI: 0.08, 0.32; depressive symptoms interaction effect = 1.84, 95% CI: 0.44, 3.24).

**Figure 1.**
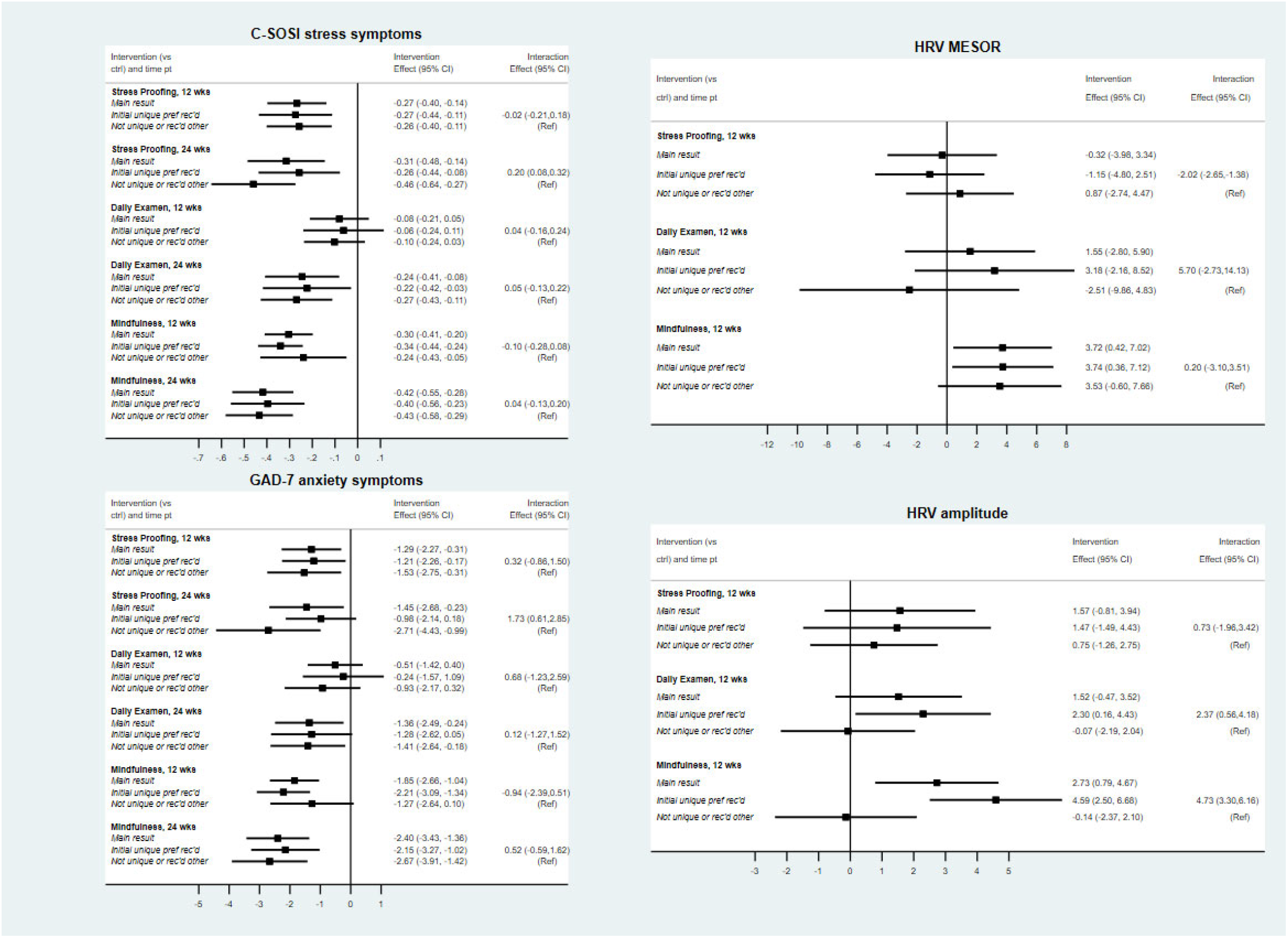
Subgroup analysis of heterogeneity of treatment effects by preference type

### 3.8 Sensitivity Analysis

When missing outcome and covariate values were imputed using MICE methods, results for stress, anxiety, and depressive symptoms at 12 weeks did not differ substantially from complete case estimates (Supplemental Table B9). However, magnitudes of treatment effects at 24 weeks were attenuated when missing values were imputed, suggesting that participants with lower stress, anxiety, and depressive symptom scores may have been more likely to drop out of the study between 12 and 24 weeks. In contrast, multiply imputed estimates for HRV outcomes moved in the direction of better HRV outcomes across all interventions.

Results of the trial data were largely similar to pooled results with observational and post-waitlist data (Supplemental Table B10).

## 4 DISCUSSION

We performed a partially randomized, patient-preference, waitlist control study to evaluate the effectiveness of three potentially stress-reducing interventions on self-reported symptoms of stress and one biological marker of parasympathetic nervous system activity among clergy in North Carolina. Modality of delivery of interventions was modified from in-person to online to accommodate for changes imposed by COVID-19. We analyzed data from 255 participants who underwent randomization.

Separate profiles of evidence emerged when each intervention was independently compared to the waitlist control. Participants allocated to MBSR evidenced improvement in self-reported symptoms of stress, anxiety and depressed mood, and in HRV MESOR and amplitude from pre-to post-intervention at 12 weeks, with improvements in symptoms of stress, anxiety, and depressed mood maintained at 24 weeks. Participants allocated to Stress Proofing evidenced improvements in symptoms of stress, anxiety, and depressed mood from pre-to post-intervention at 12 weeks and maintained at 24 weeks but did not evidence change in HRV parameters. Improvement in symptoms of stress, anxiety, and depressed mood among participants who completed the Daily Examen were not evidenced until 24 weeks with no change evidenced in HRV parameters. Stated alternately, participation in MBSR resulted in stable and enduring improvement in self-reported and physiological correlates of stress, while participation in Stress Proofing resulted in enduring improvement in self-reported correlates of stress, and participation in the Daily Examen resulted in delayed improvements in self-reported correlates of stress.

We included MBSR as a gold standard stress management intervention. The enduring improvements in symptoms observed among participants allocated to MBSR are consistent with a systematic review of the effects of MBSR interventions among non-clinical samples that reported significant improvements in symptoms of stress, anxiety, and depression when compared to nonactive control conditions [63]. In the current study, these findings may be attributed to high engagement; participants who reported practice on a given day averaged 30 minutes, which is consistent with averages across mindfulness-based interventions [19] and can create clinical changes. For example, thirty minutes of daily MBSR practice increases gray matter concentration in brain regions important in emotion regulation [64].

We hypothesized that the Daily Examen may influence symptoms of stress through mechanisms are similar to those observed in MBSR [65]. Both practices develop the ability to observe and describe thoughts, feelings, and behaviors, which may help bring attention to the present (or past 24 hours) as opposed to worry about the future. Further, both practices encourage non-reactivity toward thoughts and feelings, which may promote calmness. Statistically significant improvements in self-reported correlates of stress were not evidenced among participants allocated to the Daily Examen until 24 weeks, suggesting lagged effects relative to MBSR and that it may benefit from increased practice duration or frequency. Nevertheless, among participants who provided engagement data, Daily Examen practice was high throughout the 24 weeks. A prayer practice may be particularly acceptable for populations such as clergy and Christians generally. The Daily Examen may be more feasible than MBSR to sustain past 24 weeks at just 15 minutes of practice per day. Only one empirical study on the Daily Examen other than our pilot study has been published focusing on positive emotions; participants randomly assigned to practice the Examen increased in self-transcendent positive emotions but not eudaimonic motivation after two weeks [66]. Thus, the current study is the first to empirically investigate the effects of the Daily Examen on stress outcomes.

Consistent with studies on stress inoculation therapy [67, 20], Stress Proofing -- a set of stress inoculation, breathing, and walking exercises plus lifestyle changes -- led to post-intervention and enduring improvements in self-reported correlates of stress. Stress Proofing exercises differed from mindfulness-based exercises in that participants were not explicitly taught to direct their thoughts to the present. For example, in the MBSR awareness of breath exercise, participants were taught to notice their breath without changing it as a way to focus on the present, whereas in Stress Proofing, participants were taught to change their breathing (e.g., triangle, square, and deep breathing) without intentional present focus. The goal of the Stress Proofing breathing exercises was to lower heart rate and impact the autonomic nervous system; other studies have found that diaphragmatic breathing decreases diastolic and systolic blood pressure, salivary cortisol, respiratory rate, and anxiety symptoms, although researchers have called for more high-quality studies to determine clinical utility [67, 68]. Stress Proofing did not evidence improvement in HRV at 12 weeks despite the physical focus of Stress Proofing practices. Like the Daily Examen, suggested daily practice for Stress Proofing was 15 minutes.

Only MBSR participants evidenced a statistically significant improvement in HRV, a noninvasive biological marker of the strength of the parasympathetic nervous system as measured at the sinoatrial node [69]. Following completion of MBSR, participants evidenced improvement in two circadian HRV parameters: the MESOR which reflects trait-like activity of the parasympathetic nervous system, and amplitude which reflects higher day-to-day variability of parasympathetically-mediated HRV. Previous research evaluating the effect of MBSR on HRV is equivocal, with one systematic review that identified 19 randomized controlled trials (RCTs) evaluating the effect of mindfulness-based interventions on HRV, reporting no statistically significant difference pre- to post-intervention (Hedges’ g = 0.38, 95% CI = - 0.014 to 0.77) [36]. It is important to note that there were only two RCTs included in this review that evaluated MBSR and included 24-hour HRV as an outcome, one within 168 people who lived with fibromyalgia [70] and another within 19 people who experienced benign heart palpitations [71]. As such, the current trial provides the most robust evidence to date for the effect of MBSR on long-term HRV when delivered with fidelity among a community sample.

Confidence in the effect of MBSR on HRV observed in the present study is strengthened for five reasons: 1) propensity score adjustment was performed to ensure that results could not be explained by variation in baseline characteristics associated with HRV, such as age and gender [72], body mass index [73], and physical activity [74]; 2) HRV was quantified using long-term records and time-domain analysis which are less sensitive to transient influences, such as change in respiration and movement; 3) the effect of MBSR on HRV was observed in the trial sample as well as the observational sample and was strengthened following imputation of missing data; 4) MBSR was delivered with fidelity by [removed for blind review], and 5) practice data received indicated high engagement. While we can conclude that MBSR resulted in improvement in parasympathetic cardiac control with good confidence, no measure of sympathetic activation was collected (e.g., pre-ejection period), and conclusions cannot be made about the effect of MBSR on activation of the sympathetic nervous system.

The Selah study is one of few to conduct a behavioral trial of a specific prayer practice. The Daily Examen was preferred by more participants than the other two interventions, although it had attenuated effects even at 24 weeks. However, given the degree of acceptability shown across interventions, it may be possible to increase the amount of prayer time or combine the Daily Examen with elements of MBSR to lead to physiological benefit, although this would require further testing.

In this trial, there was clear engagement for all three practices during the initial three-month intervention period and persisting for an additional three months. Such engagement could have been driven by a need for stress symptom reduction due to the COVID-19 pandemic, or because the practices were feasible and acceptable even for busy clergy professionals. The interventions tested in this study conveyed benefits strong enough to surface during this period of uncertainty with its multiple and shifting stressors.

Results largely did not support our hypothesis that participants who received their preferred intervention would evidence larger improvement in outcomes of interest compared to waitlist control. Outcomes of interest were only observed to vary by preference among two comparisons. Participants allocated to Stress Proofing who did not receive their initially preferred intervention observed greater reduction in symptoms of stress at 24 weeks, suggesting greater durability of effects when a unique preference was not present. The observation that outcomes of interest were largely unaffected by preference may be explained by the high rate of practice and the large portion of the sample who received their preferred intervention (∼80% of trial participants).

This study had several limitations. We did not conduct intervention fidelity checks, although this concern is somewhat tempered knowing that interventions were delivered by certified MBSR instructors and all instructors followed the same materials throughout the trial. HRV data was collected at 12 but not 24 weeks, which limits our understanding of the durability of treatment effects on HRV parameters. HRV data collected at 24 weeks would have been particularly interesting for the Daily Examen intervention which saw improvements in self-reported correlates of stress at 24 weeks and not at 12 weeks.

Observed results are limited to individuals in a single profession (clergy from a single, predominantly white denomination) in a single geographic area. We operationalized stress as a response within the current trial. In other words, our primary outcomes included physical and psychological responses that are typically associated with stress (e.g., symptoms of anger, muscle tension, cardiopulmonary arousal), and a biological marker that is an integral part of the stress response system with important implications for stress and health [30]. These outcomes are temporally removed from acute stressors and believed to indirectly approximate neurobiological changes in response to daily stressors experienced over the course of weeks [74]. Importantly, such measures fail to adequately capture exposures to stressors, cognitive appraisals associated with stressors, or real or perceived abilities to cope with stress [75]. We acknowledge that the changes observed in our primary outcomes of interest may be due to influences other than changes in the way participants appraise or cope with stressors (i.e., the exact mechanisms explaining differences observed are not known).

In addition, although partially randomized preference trials may enhance external validity of a study [75], estimates of treatment effects may suffer from similar biases as may be seen in an observational study due to confounding between characteristics that give rise to preference and the outcome under study. Some previous studies suggest that analyzing outcomes of a study on the subsample of participants who were indifferent to their intervention allocation can provide an unbiased treatment effect estimate [76]. Our sample size did not allow for this analysis, the results of which may have added confidence to our findings. Another approach – used in the current study – is to control for confounding characteristics between preference and the outcome, which may lead to estimates with more precision and a relatively unbiased estimate of the treatment effect [77]. However, the propensity score adjustment approach can only correct confounding bias if the propensity model is correctly specified [55], which cannot be definitively confirmed. Further, treatment assignment and partial randomization occurred prior to baseline data collection which limited our ability to characterize participants who withdrew from the study and leaves the possibility that randomization may have affected the baseline level of outcomes. Finally, the partially randomized structure of intervention assignment means there were limited cases in which a participant received an intervention that they specifically did not initially prefer, thus we could not explicitly measure the effect of preference on study outcomes, only a selection effect [78].

This study also had several strengths. We evaluated both emerging and well-validated interventions and did so using a partially randomized preference design that accounted for participants’ preferences, which in behavioral trials have the potential to affect engagement and outcomes. Throughout the trial, we used the same instructors for Daily Examen and Stress Proofing, and all of the MBSR instructors met well-established MBSR certification standards. The trial methods were adapted to COVID-19 in a way that approximated real-world conditions, and practice was measured for 24 weeks with high daily response rates. We collected self-report and physiological measures for a relatively large sample. HRV is not subject to expectancy effects and provides confidence in the improvements seen in the MBSR participants, and simultaneously, the self-reported symptom outcomes are useful in indicating that participants across interventions felt noticeably better, even if it was a placebo effect. The fact that Daily Examen participants did not have significantly improved scores across a variety of self-reported outcomes until 24 months also provides some confidence in the timing of feeling better for each intervention for the average participant. Further, we were able to use existing survey data from the study population to describe participants who selected into the trial, informing generalizability and possible bias.

## 5 CONCLUSION

The Selah Stress Management Trial tested three separate behavioral interventions compared to a control group. Participants who provided text message data engaged in each intervention with great frequency and with enduring practice through a 12-week intervention and an additional 12 weeks, indicating that each intervention was acceptable to clergy who are busy professionals who engage in emotional and administrative activities. Each intervention group experienced improvements in self-reported correlates of stress at 24 weeks, during a particularly stressful time of the COVID-19 pandemic, and with two of the interventions requiring only 15 minutes of practice per day. Only MBSR, which, when practiced and reported, was practiced on average 30 minutes per day, resulted in statistically significant improvement in HRV from pre-to post-intervention. Despite a robust literature of MBSR’s effects on self-reported correlates of stress, this is the first study to show a significant improvement on circadian HRV parameters. These findings show the strongest evidence of improvement for MBSR, although Stress Proofing and the Daily Examen may be considered if individuals do not prefer MBSR. There is a clear need for stress symptom reduction among many occupational groups; these findings provide evidence of effectiveness of three manageable and scalable interventions.

## Supporting information

Supplement A B CONSORT

## Data Availability

The datasets generated during the current study are not publicly available but we will make de-identified data available for reasonable requests compliant with ethical approvals from the sending and receiving hosts institutional ethics review boards.

## ACKNOWLEDGMENTS

The authors greatly appreciate the numerous instructors who designed and implemented the interventions: Glenn Murphy, Mark Shaw, Karen Keen, Riitta Whaley, Maya McNeilly, Phyllis Hicks, and Julie Kosey. For her recruitment efforts, we thank Claire Cusick. For interview, text message, and heart rate variability data collection, we thank Beth Stringfield, Blen Biru, Jessica Choi, and Sofia Labrecque, and for data collection and heart rate variability processing, we thank Georgia Price, Grace Geib, James Boyd, Jennie Sun, Malcolm Smith Fraser, Meagan Gillette, Sean Kehoe, Grace Smith, Khue Huynh, Andrew Takais, Bentley Choi, David Steele, Emily Woodrow, Emmy Duerr, Lori Parrish, Sarah Moninger, Maya Montgomery, Sofia Labrecque, and Ben Westlund. We thank Elizabeth Wallack for processing and quantification of heart rate variability data files. We appreciate consultations with Liz Turner and Joe Egger of the Duke Global Health Institute Research Design and Analysis Core and Carl Weisner of Duke Divinity School, literature review from Celia Hybels and Anna Holleman, and manuscript comments from the Clergy Health Initiative Writing Group.

## ACRONYMS

(HRV): Mindfulness-Based Stress Reduction (MBSR) Heart rate variability
(UMC): United Methodist Church
(C-SOSI): Calgary-Symptoms of Stress Inventory
(MESOR): Midline Estimating Statistic Of Rhythm
(GAD-7): Generalized Anxiety Disorder-7
(PHQ-8): Patient Health Questionnaire-8
(ICC): Interclass correlation coefficient
(cLDA): Constrained longitudinal data analysis modeling
(MICE): Multiple imputation with chained equations
(ms): Milliseconds
(MD): Mean difference
(BMI): Body Mass Index
(RCTs): Randomized controlled trials

## REFERENCES

1. Cohen S, Gianaros PJ, Manuck SB. A stage model of stress and disease. Perspect Psychol Sci. 2016 Jul;11(4):456–63.

2. Lovallo WR. Stress and health: biological and psychological interactions. SAGE Publications; 2015. 353 p.

3. Kivimäki M, Steptoe A. Effects of stress on the development and progression of cardiovascular disease. Nat Rev Cardiol. 2018 Apr;15(4):215–29.

4. Richardson S, Shaffer JA, Falzon L, Krupka D, Davidson KW, Edmondson D. Meta-analysis of perceived stress and its association with incident coronary heart disease. Am J Cardiol. 2012 Dec 15;110(12):1711–6.

5. Cohen S, Janicki-Deverts D, Miller GE. Psychological stress and disease. JAMA. 2007 Oct 10;298(14):1685–7.

6. Proeschold-Bell RJ, LeGrand SH. High rates of obesity and chronic disease among United Methodist clergy. Obesity. 2010;18(9):1867–70.

7. Webb BL, Chase K. Occupational distress and health among a sample of Christian clergy. Pastoral Psychology. 2019;68:331–43.

8. Mook AM. Prevalence of chronic disease, associated factors, and health related quality of life among Wesleyan clergy [dissertation]. United States -- Indiana: Indiana State University; 2019.

9. Lindholm G, Johnston J, Dong F, Moore K, Ablah E. Clergy Wellness: An assessment of perceived barriers to achieving healthier lifestyles. J Relig Health. 2016 Feb;55(1):97–109.

10. Proeschold-Bell RJ, Miles A, Toth M, Adams C, Smith BW, Toole D. Using effort-reward imbalance theory to understand high rates of depression and anxiety among clergy. J Primary Prevent. 2013 Dec 1;34(6):439–53.

11. Jones SH, Francis LJ, Jackson C. The relationship between religion and anxiety: A study among Anglican clergymen and clergywomen. Journal of Psychology and Theology. 2004 Jun 1;32(2):137–42.

12. Weaver AJ, Larson DB, Flannelly KJ, Stapleton CL, Koenig HG. Mental health Issues among clergy and other religious professionals: A review of research. J Pastoral Care Counsel. 2002 Dec 1;56(4):393–403.

13. Knox S, Virginia SG, Thull J, Lombardo JP. Depression and contributors to vocational satisfaction in Roman Catholic secular clergy. Pastoral Psychol. 2005 Nov 1;54(2):139–55.

14. Varvogli L, Darviri C. Stress management techniques: evidence-based procedures that reduce stress and promote health. Health Science Journal. 2011;5(2).

15. Sedlmeier P, Eberth J, Schwarz M, Zimmermann D, Haarig F, Jaeger S, et al. The psychological effects of meditation: A meta-analysis. Psychological Bulletin. 2012;138:1139–71.

16. Janssen M, Heerkens Y, Kuijer W, van der Heijden B, Engels J. Effects of mindfulness-based stress reduction on employees’ mental health: A systematic review. PLoS One. 2018 Jan 24;13(1):e0191332.

17. Kriakous SA, Elliott KA, Lamers C, Owen R. The effectiveness of mindfulness-based stress reduction on the psychological functioning of healthcare professionals: a systematic review. Mindfulness (N Y). 2021;12(1):1–28.

18. Abbott RA, Whear R, Rodgers LR, Bethel A, Thompson Coon J, Kuyken W, et al. Effectiveness of mindfulness-based stress reduction and mindfulness based cognitive therapy in vascular disease: A systematic review and meta-analysis of randomised controlled trials. Journal of Psychosomatic Research. 2014 May 1;76(5):341–51.

19. Parsons CE, Crane C, Parsons LJ, Fjorback LO, Kuyken W. Home practice in mindfulness-based cognitive therapy and mindfulness-based stress reduction: A systematic review and meta-analysis of participants’ mindfulness practice and its association with outcomes. Behav Res Ther. 2017 Aug;95:29–41.

20. Saunders T, Driskell JE, Johnston JH, Salas E. The effect of stress inoculation training on anxiety and performance. Journal of Occupational Health Psychology. 1996;1:170–86.

21. Kashani F, Kashani P, Moghimian M, Shakour M. Effect of stress inoculation training on the levels of stress, anxiety, and depression in cancer patients. Iran J Nurs Midwifery Res. 2015;20(3):359–64.

22. Hofmann SG, Asnaani A, Vonk IJJ, Sawyer AT, Fang A. The efficacy of cognitive behavioral therapy: A review of meta-analyses. Cognit Ther Res. 2012 Oct 1;36(5):427–40.

23. Butler AC, Chapman JE, Forman EM, Beck AT. The empirical status of cognitive behavioral therapy: A review of meta-analyses. Clinical Psychology Review. 2006 Jan 1;26(1):17–31.

24. Lindsay EK, Creswell JD. Mechanisms of mindfulness training: Monitor and Acceptance theory (MAT). Clin Psychol Rev 2017;51:48–59.

25. Meichenbaum, D. Stress inoculation training: A preventative and treatment approach. 3^rd^ ed. Lehrer PM, Sime WE, editors. New York: Guilford Press; 2005.

26. Meichenbaum, D., Cameron, R. Stress inoculation training: Toward a general paradigm for training coping skills. Meichenbaum D, Jaremko ME, editors. Boston (MA): Spring; 1983. p. 115–154.

27. Crosswell AD, Lockwood KG. Best practices for stress measurement: How to measure psychological stress in health research. Health Psychol Open. 2020 Jul 8;7(2):2055102920933072.

28. Thayer JF, Åhs F, Fredrikson M, Sollers JJ, Wager TD. A meta-analysis of heart rate variability and neuroimaging studies: Implications for heart rate variability as a marker of stress and health. Neuroscience & Biobehavioral Reviews. 2012 Feb 1;36(2):747–56.

29. Kim HG, Cheon EJ, Bai DS, Lee YH, Koo BH. Stress and heart rate variability: A meta-analysis and review of the literature. Psychiatry Investig. 2018 Mar;15(3):235–45.

30. Kemp AH, Quintana DS, Felmingham KL, Matthews S, Jelinek HF. Depression, comorbid anxiety disorders, and heart rate variability in physically healthy, unmedicated patients: Implications for cardiovascular risk. PLoS One. 2012 Feb 15;7(2):e30777.

31. Kemp AH, Quintana DS, Gray MA, Felmingham KL, Brown K, Gatt JM. Impact of depression and antidepressant treatment on heart rate variability: A review and meta-analysis. Biological Psychiatry. 2010 Jun 1;67(11):1067–74.

32. Kemp AH, Brunoni AR, Santos IS, Nunes MA, Dantas EM, Carvalho de Figueiredo R, et al. Effects of depression, anxiety, comorbidity, and antidepressants on resting-state heart rate and its variability: an ELSA-Brasil cohort baseline study. Am J Psychiatry. 2014 Dec 1;171(12):1328–34.

33. Järvelin-Pasanen S, Sinikallio S, Tarvainen MP. Heart rate variability and occupational stress-systematic review. Ind Health. 2018 Nov 21;56(6):500–11.

34. Liao D, Carnethon M, Evans GW, Cascio WE, Heiss G. Lower heart rate variability is associated with the development of coronary heart disease in individuals with diabetes: the atherosclerosis risk in communities (ARIC) study. Diabetes. 2002 Dec;51(12):3524–31.

35. Tsuji H, Larson MG, Venditti FJ, Manders ES, Evans JC, Feldman CL, et al. Impact of reduced heart rate variability on risk for cardiac events. The Framingham Heart Study. Circulation. 1996 Dec 1;94(11):2850–5.

36. Brown L, Rando AA, Eichel K, Van Dam NT, Celano CM, Huffman JC, et al. The effects of mindfulness and meditation on vagally mediated Heart Rate Variability: A meta-Analysis. Psychosom Med. 2021 Aug 1;83(6):631–40.

37. Walter SD, Turner R, Macaskill P, McCaffery KJ, Irwig L. Beyond the treatment effect: Evaluating the effects of patient preferences in randomised trials. Stat Methods Med Res. 2017 Feb;26(1):489–507.

38. Sidani S, Epstein DR, Bootzin RR, Moritz P, Miranda J. Assessment of preferences for treatment: validation of a measure. Res Nurs Health. 2009 Aug;32(4):419–31.

39. StataCorp. 2019. Stata Statistical Software: Release 16. College Station, TX: StataCorp LLC.

40. Van Dam NT, van Vugt MK, Vago DR, Schmalzl L, Saron CD, Olendzki A, et al. Mind the hype: A critical evaluation and prescriptive agenda for research on mindfulness and meditation. Perspect Psychol Sci. 2018 Jan;13(1):36–61.

41. Kabat-Zinn, J. Meditation is not what you think: mindfulness and why it is so important. 1st ed. New York: Hachette Books; 2018.

42. Thibodeaux SJ ME. Reimagining the Ignatian examen: fresh ways to pray from your day. Loyola Press; 2015.

43. Lazarus RS, Folkman S. Stress, appraisal, and coping. Springer Publishing Company; 1984. 460 p.

44. Carlson LE, Thomas BC. Development of the Calgary Symptoms of Stress Inventory (C-SOSI). Int J Behav Med. 2007;14(4):249–56.

45. Penwell, Lauren M., “Validation of the Calgary Symptoms of Stress Inventory (C-SOSI) for Predicting adherence to a stress reduction technique” (2012). Graduate Theses, Dissertations, and Problem Reports. 4909.

46. Johnson SU, Ulvenes PG, Øktedalen T, Hoffart A. Psychometric properties of the General Anxiety Disorder 7-Item (GAD-7) scale in a heterogeneous psychiatric sample. Front Psychol. 2019 Aug 6;10:1713.

47. Plummer F, Manea L, Trepel D, McMillan D. Screening for anxiety disorders with the GAD-7 and GAD-2: a systematic review and diagnostic meta-analysis. Gen Hosp Psychiatry. 2016 Mar-Apr;39:24–31. doi: 10.1016/j.genhosppsych.2015.11.005. Epub 2015 Nov 18. PMID: 26719105.

48. Kroenke K, Strine TW, Spitzer RL, Williams JBW, Berry JT, Mokdad AH. The PHQ-8 as a measure of current depression in the general population. J Affect Disord. 2009 Apr;114(1–3):163–73.

49. Kroenke K, Spitzer RL, Williams JBW, Löwe B. The Patient Health Questionnaire Somatic, Anxiety, and Depressive Symptom Scales: A systematic review. Gen Hosp Psychiatry. 2010;32(4):345–59.

50. Sidani S, Epstein DR, Bootzin RR, Moritz P, Miranda J. Assessment of preferences for treatment: validation of a measure. Res Nurs Health. 2009; 32(4):419–31. https://doi.org/10.1002/nur.20329.

51. PA Harris, R Taylor, BL Minor, V Elliott, M Fernandez, L O’Neal, L McLeod, G Delacqua, F Delacqua, J Kirby, SN Duda, REDCap Consortium, The REDCap consortium: Building an international community of software partners, J Biomed Inform. 2019 May 9 [doi: 10.1016/j.jbi.2019.103208]

52. Tarvainen MP, Niskanen J-P, Lipponen JA, Ranta-Aho PO, Karjalainen PA. Kubios HRV--heart rate variability analysis software. Comput Methods Programs Biomed. 2014;113(1):210–20

53. Twilio, Inc. 2020. Twilio Customer Engagement Platform. https://www.twilio.com/docs/api

54. Laborde S, Mosley E, Thayer JF. Heart rate variability and cardiac vagal tone in psychophysiological research: Recommendations for experiment planning, data analysis, and data reporting. Front Psychol. 2017;8:213.

55. Vansteelandt S, Daniel R m. On regression adjustment for the propensity score. Statistics in Medicine. 2014;33(23):4053–72.

56. Imai K, Ratkovic M. Covariate balancing propensity score. J R Stat Soc B. 2014 Jan;76(1):243–63.

57. Flight L, Allison A, Dimairo M, Lee E, Mandefield L, Walters SJ. Recommendations for the analysis of individually randomised controlled trials with clustering in one arm - a case of continuous outcomes. BMC Med Res Methodol. 2016 Nov 29;16(1):165.

58. Coffman CJ, Edelman D, Woolson RF. To condition or not condition? Analysing “change” in longitudinal randomised controlled trials. BMJ Open. 2016 Dec 30;6(12):e013096.

59. White IR, Royston P, Wood AM. Multiple imputation using chained equations: Issues and guidance for practice. Stat Med. 2011 Feb 20;30(4):377–99.

60. Rubin DB. Multiple Imputation for Nonresponse in Surveys. Subsequent. Hoboken, N.J: Wiley-Interscience; 2004. p. 258.

61. Rubin M. When to adjust alpha during multiple testing: a consideration of disjunction, conjunction, and individual testing. Synthese. 2021 Dec 1;199(3):10969–1000.

62. Benjamini Y, Yekutieli D. The control of the false discovery rate in multiple testing under dependency. The Annals of Statistics. 2001 Aug;29(4):1165–88.

63. Querstret D, Morison L, Dickinson S, Cropley M, John M. Mindfulness-based stress reduction and mindfulness-based cognitive therapy for psychological health and well-being in nonclinical samples: A systematic review and meta-analysis. International Journal of Stress Management. 2020;27:394–411.

64. Hölzel BK, Carmody J, Vangel M, Congleton C, Yerramsetti SM, Gard T, et al. Mindfulness practice leads to increases in regional brain gray matter density. Psychiatry Res. 2011 Jan 30;191(1):36–43.

65. Alsubaie M, Abbott R, Dunn B, Dickens C, Keil TF, Henley W, et al. Mechanisms of action in mindfulness-based cognitive therapy (MBCT) and mindfulness-based stress reduction (MBSR) in people with physical and/or psychological conditions: A systematic review. Clin Psychol Rev. 2017 Jul;55:74–91.

66. Curlee MS, Ahrens AH. An exploratory analysis of the Ignatian examen: Impact on self-transcendent positive emotions and eudaimonic motivation. The Journal of Positive Psychology. 2022 Aug 7;0(0):1–10.

67. Hopper SI, Murray SL, Ferrara LR, Singleton JK. Effectiveness of diaphragmatic breathing for reducing physiological and psychological stress in adults: a quantitative systematic review. JBI Database System Rev Implement Rep. 2019 Sep;17(9):1855–76.

68. Hamasaki H. Effects of diaphragmatic breathing on health: A narrative review. Medicines (Basel). 2020 Oct 15;7(10):65.

69. Berntson GG, Bigger JT, Eckberg DL, Grossman P, Kaufmann PG, Malik M, et al. Heart rate variability: origins, methods, and interpretive caveats. Psychophysiology. 1997 Nov;34(6):623–48.

70. Grossman P, Deuring G, Walach H, Schwarzer B, Schmidt S. Mindfulness-based intervention does not influence cardiac autonomic control or the pattern of physical activity in fibromyalgia during daily life: An ambulatory, multi-measure randomized controlled trial. Clin J Pain. 2017 May;33(5):385–94.

71. Plews-Ogan M, Owens JE, Goodman M, Wolfe P, Schorling J. A pilot study evaluating mindfulness-based stress reduction and massage for the management of chronic pain. J Gen Intern Med. 2005 Dec;20(12):1136–8.

72. Umetani K, Singer DH, McCraty R, Atkinson M. Twenty-four hour time domain heart rate variability and heart rate: relations to age and gender over nine decades. J Am Coll Cardiol. 1998 Mar 1;31(3):593–601.

73. Yi SH, Lee K, Shin DG, Kim JS, Kim HC. Differential association of adiposity measures with heart rate variability measures in Koreans. Yonsei Med J. 2013 Jan 1;54(1):55–61.

74. Soares-Miranda L, Sattelmair J, Chaves P, Duncan G, Siscovick DS, Stein PK, et al. Physical activity and heart rate variability in older adults: The cardiovascular health study. Circulation. 2014 May 27;129(21):2100–10.

75. Epel ES, Crosswell AD, Mayer SE, Prather AA, Slavich GM, Puterman E, et al. More than a feeling: A unified view of stress measurement for population science. Front Neuroendocrinol. 2018 Apr;49:146–69.

76. Crosswell AD, Epel ES, Mendes WB, Prather AA. Improving the language specificity of stress in psychological and population health science. Psychosom Med. 2022 Jun 1;84(5):643–4.

77. TenHave TR, Coyne J, Salzer M, Katz I. Research to improve the quality of care for depression: alternatives to the simple randomized clinical trial. Gen Hosp Psychiatry. 2003;25(2):115–23.

78. Walter SD, Bian M. Relative efficiencies of alternative preference-based designs for randomised trials. Stat Methods Med Res. 2020 Dec;29(12):3783–803.

79. Gemmell I, Dunn G. The statistical pitfalls of the partially randomized preference design in non-blinded trials of psychological interventions. Int J Methods Psychiatr Res. 2011 Feb 24;20(1):1–9.

