## Supplement A B CONSORT for "The Selah trial: A preference-based partially randomized waitlist control study of three stress management interventions"

### Appendix A. Supplemental Methods

**Study Hypotheses**

Hypothesis 1a: Compared to the waitlist control condition, stress symptoms will be relieved at 12 weeks for MBSR, the Daily Examen, and Stress Proofing when evaluated independently.

Hypothesis 1b: Compared to the waitlist control condition, stress symptoms will be relieved at 24 weeks for MBSR, the Daily Examen, and Stress Proofing when evaluated independently.

Hypothesis 2a: Compared to the waitlist control condition, anxiety symptoms will be relieved at 12 weeks for MBSR, the Daily Examen, and Stress Proofing when evaluated independently.

Hypothesis 2b: Compared to the waitlist control condition, anxiety symptoms will be relieved at 24 weeks for MBSR, the Daily Examen, and Stress Proofing when evaluated independently.

Hypothesis 3a: Compared to the waitlist control condition, depression symptoms will be relieved at 12 weeks for MBSR, the Daily Examen, and Stress Proofing when evaluated independently (exploratory outcome).

Hypothesis 3b: Compared to the waitlist control condition, depression symptoms will be relieved at 24 weeks for MBSR, the Daily Examen, and Stress Proofing when evaluated independently (exploratory outcome).

Hypothesis 4: Compared to the waitlist control condition, HRV will be improved at 12 weeks for MBSR, the Daily Examen, and Stress Proofing when evaluated independently.

Hypotheses 5a-5d: Participants who had a stated preference and received that intervention (i.e. MBSR, the Daily Examen, and Stress Proofing combined) will experience larger between-arm (waitlist vs non-waitlist) differences in improvements on a) stress symptoms at 12 weeks; b) anxiety symptoms at 12 weeks; c) depression symptoms at 12 weeks; and d) HRV at 12 weeks, when compared to no-preference participants randomly assigned across interventions and waitlist (exploratory outcome).

Hypotheses 5e-5g: Participants who had a stated preference and received that intervention (i.e. MBSR, the Daily Examen, and Stress Proofing combined) will experience larger between-arm (waitlist vs non-waitlist) differences in improvements on e) stress symptoms at 24 weeks, f) anxiety symptoms at 24 weeks, and g) depression symptoms at 24 weeks, when compared to no-preference participants randomly assigned across interventions and waitlist (exploratory outcome).

**Supplemental Methods Figure A1.** Pandemic-adapted Selah study design: A partially-randomized waitlist-controlled preference trial design

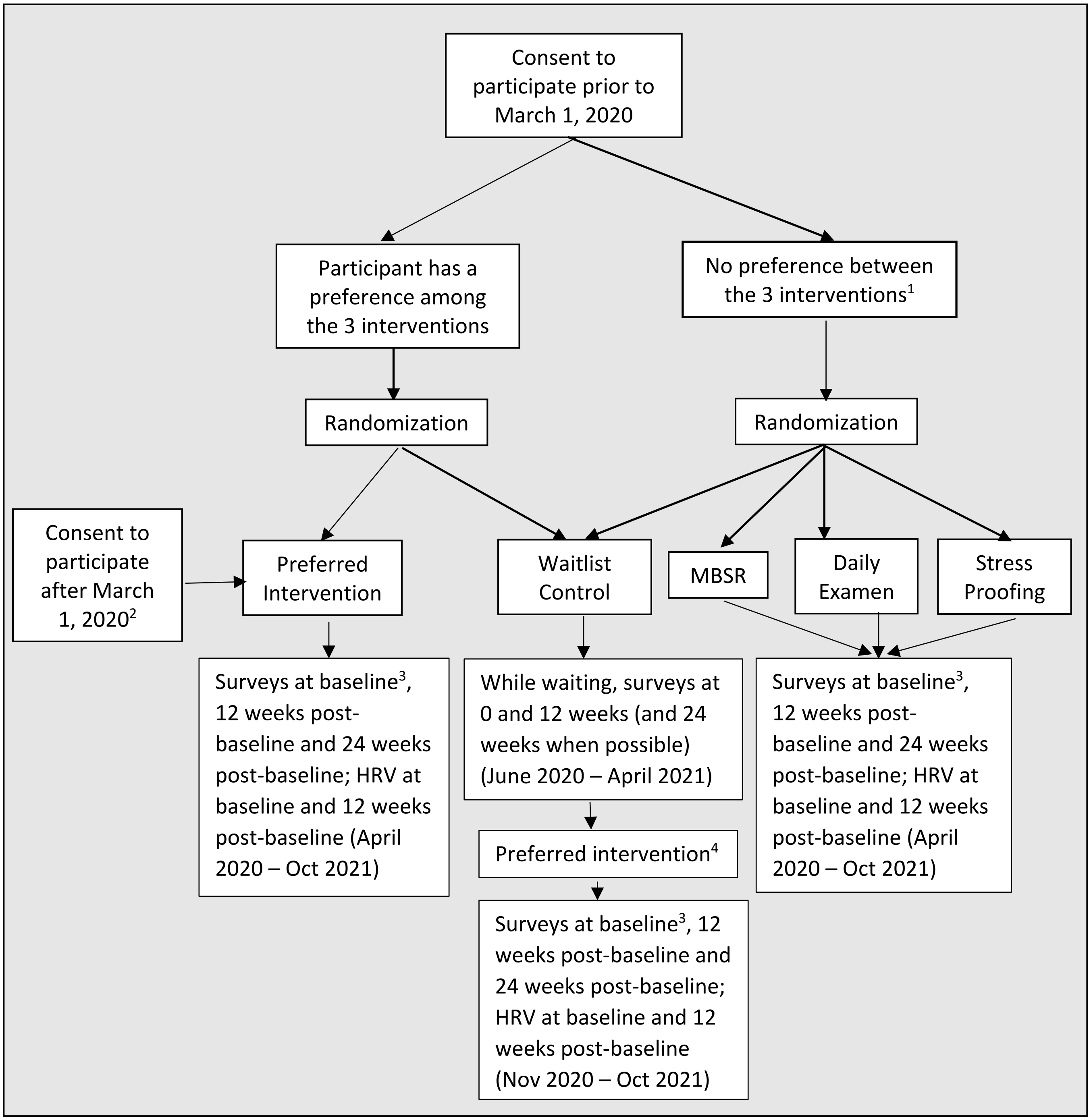


[1] Participants who prefer 2 interventions equally and over the third will be first randomly assigned between the 2 preferred interventions, and then randomized into either their preferred intervention or waitlist control group.
[2] All participants who consented after March 1, 2020 were non-randomly assigned to their preferred intervention. Participants with no preference selected a workshop with dates of their choosing.

[3] Baseline indicates immediately before intervention start.
[4] After waitlist control participants finish giving control data, they may participate in the intervention they originally indicated as their preference or change it to reflect their current preference.

**Supplemental Methods Table A1**. Study assignment and allocation approach for each preference and enrolment date scenario

| Preference Scenario | Assignment Approach |
|---|---|
| **Enrolled prior to March 1, 2020** | |
| No preference between interventions | Randomly assigned to one of the four study arms: three interventions without waitlist and the waitlist arm, with a 1:1:1:1 ratio |
| Preferred two interventions equally and over the third intervention | Randomly assigned to one of the two interventions with a 1:1 ratio, and then a fraction was randomly assigned to the waitlist arm, with a 3:1 non-waitlist vs waitlist ratio for MBSR and Stress Proofing and a 5:4 non-waitlist vs waitlist ratio for the Daily Examen (DE was preferred by more participants and this allowed the same number of participants to be randomized into each study arm) |
| Preferred one intervention among the three | Assigned to their preferred intervention and combined with participants with two top preferences who had been randomized to that intervention, and then randomly assigned to non-waitlist vs waitlist arms, with a 3:1 non-waitlist vs waitlist ratio for MBSR and Stress Proofing and a 5:4 non-waitlist vs waitlist ratio for the Daily Examen |
| Any of the above scenarios and are part of a married (or cohabitating) couple who both meet study criteria and enrolled | To avoid spillover effects, each couple was treated as if they were one person, i.e., assigning both spouses to the same intervention and randomizing them into a non-waitlist vs waitlist arm. When a couple had different preferences, one preference was randomly chosen as the couple's preference. |
| Seven clergy with an established meeting group (a covenant group) | The seven clergy jointly chose a single preferred intervention and were randomized together to the non-waitlist vs waitlist arm. |
| **Enrolled after March 1, 2020** | |
| All enrollees after March 1, 2020 with no preference, two equal preferences, or one preference, and whether a clergy couple or not | Participants answered treatment preference survey items but regardless of their answers, self-selected the intervention with intervention dates they most wanted from the full list of workshop options. None were randomized to the non-waitlist vs waitlist arms; they all were assigned to non-waitlist. |

**Heart Rate Variability (HRV) data collection procedures and processing**

During the enrollment process, participants were asked survey questions to determine their eligibility for inclusion in HRV data collection. Participants were excluded from HRV data collection if they had underlying medical conditions, including a diagnosis of tachycardia; being pregnant or becoming pregnant during the course of data collection; being diagnosed with COVID-19; having a pacemaker; and documentation of other cardiovascular-related chronic or acute morbidities that could impact the integrity of HRV data (Supplemental Table A4).

Two weeks prior to the intervention, participants were mailed a box with a Bittium eMotion Faros 180 recording device with electrodes. Participants were asked to attend an online, synchronous study orientation one week prior to the start of their workshop. During the study orientation, participants were oriented to a video and brochure that we created to convey the instructions for HRV data collection (see https://spiritedlife.org/hrv/). Participants were taught to connect the heart rate recording device's two electrode leads to two pre-gelled (Ag/AgCl) disposable Ambu BlueSensor wet-gel ECG electrodes placed beneath the right clavicle and left ribcage. Participants were instructed to wear this ambulatory heart rate monitoring device for a 48-hour period during week 0 and week 12, during which time participants proceeded with their usual work, exercise, bathing, and sleep routines.

Heart rate was measured using continuous electrocardiographic (ECG) recording sampled at a rate of 1,000 Hz and used to calculate heart rate variability. Study staff imported the 48-hour ECG recording to Kubios HRV Premium V3.4.1 software [1], partitioned it into 5-minute segments, visually inspected it to allow for manual correction of ectopic beats, detrended it, and then subjected it to Kubios' automatic artefact correction algorithm [2]. Heart rate variability was indexed using the time-domain metric Root Mean Square of Successive RR Differences (RMSSD) because it is less affected by breathing and a more suitable outcome measure in ambulatory studies than frequency-domain measures [3]. Five-minute segments across 24 hours of recording were subject to a cosinor analysis using the Cosinor package for R, based on recommendations for the detection of circadian rhythmicity [4].

**Additional details on survey measures**

Demographics measures

Survey items were included to measure ***sex*** (male/female); ***age*** (in years); ***race; ethnicity;*** self-reported ***physical, mental, and behavioral health conditions***; ***marital status;*** and having ***children living at home***. To capture work-related characteristics that may relate to stress, we measured ***appointment effort*** (i.e. full-time vs part-time appointed at UMC), ***bi-vocational status*** (i.e. having a job in addition to serving as clergy), kind of ***clergy appointment*** (i.e., serving a church vs in another capacity), ***number of congregations the study participant was appointed to***, and ***number of congregants pastored by the participant***.

Clinically relevant measures

Physical activity levels were measured using the ***Godin-Shephard Leisure-Time Physical Activity Questionnaire*** [5], a self-report measure of how often one has engaged in physical activity, measured separately for strenuous, moderate, and mild exercise, in the past seven days and for how many minutes per time. We used self-reported weight and height to assess ***body mass index*** [6]. We used single items to assess average daily ***caffeine intake***, and average weekly ***alcohol consumption***.

Stress-related measures

In addition to the Calgary-Symptoms of Stress Inventory [7], the survey asked about self-reported ***financial stress*** (How stressful is your current financial situation for you? Not at all to extremely), ***number of hours worked*** per week, and ***overall life stress level*** at study registration.

Preference measures

We included on the baseline survey an item for ***preference for online vs in-person intervention***. At the time of enrollment, we included the Treatment Acceptability and Preferences Scale [8] for each intervention. We also measured at study registration whether participants had already been ***practicing the Daily Examen or MBSR***, separately, at least 3 times each week.

The protocol paper (removed for blind review) offers details on the timing of each measure.

**Sample size**

Estimates of baseline outcome levels and expected effect sizes for C-SOSI used data from our non-randomized pilot study conducted prior to the full trial (removed for blind review). The average baseline C-SOSI score was 0.92 (SD=0.46) across all interventions, with 12-week follow-up scores of 0.7 (SD=0.58) for MBSR, 0.55 (SD=0.36) for Stress Proofing, and 0.51 (SD=0.38) for the Daily Examen.

Given an alpha of 0.0167 (based on a Bonferroni correction to hypothesis tests for the effects of three interventions on C-SOSI scores), a per-arm sample size of 40 for Daily Examen, 47 for Stress Proofing, and 195 for MBSR (which had a larger standard deviation, resulting in a larger sample size) yielded 80% power to detect a between-arm difference in means at 12 weeks of 0.22 for MBSR, 0.37 for Stress Proofing, and 0.41 for Daily Examen for a two-sample t-test with unequal variances, allowing for loss-to-follow-up of 20% and a design effect of 1.3 (corresponding to an ICC of 0.027 and average cluster size of 12) to account for clustering caused by the group-based intervention delivery. We calculated the design effect this way: $Design\ Effect = 1 + \delta(n-1)$ where $\delta$ is ICC and n is average cluster size. Note that this conservatively assumes there is clustering throughout the sample, however, we expect only partial clustering due to group treatment delivery.

Previous literature recommends defining a medium effect size for HRV as a standardized mean difference of 0.50 [9] A per-arm sample size of 140 was calculated to yield 80% power to detect an effect size of 0.50 for a two-sample t-test with an alpha of 0.0167, allowing for loss-to-follow-up of 20% and a design effect of 1.3 to account for group-based intervention delivery. We recognized that this sample size was ambitious and analysis of HRV data may lack adequate statistical power.

While preliminary published sample size calculation adjustment for multiple comparisons treated hypotheses of positive effects for each of the 3 interventions as disjunction testing (i.e. 3 tests ), upon more careful consideration the study team determined that tests for the individual interventions should be considered individual testing and testing of two separate primary outcomes (C-SOSI and HRV) should be considered disjunction testing, thus alpha adjustment occurred separately within each of the 3 interventions and adjusted for two hypotheses based on the two primary study outcomes [10].

**Propensity score methods to balance baseline characteristics between arms**

As noted in the manuscript, use of the partially randomized preference design during the trial period meant that by design the analytic data would be a mix of randomized data (for trial participants that had no preference) and observational data (for trial participants that had a preference and were allowed to select their intervention), which made it likely that treatment arms would be imbalanced on baseline characteristics in an unadjusted analysis. In addition, randomization was performed prior to baseline data collection, with substantial study dropout in the interim. Thus, statistical analysis necessitated incorporation of observational methods to rebalance the intervention arms on characteristics that may have influenced selection into a particular intervention. A propensity score covariate adjustment method [11] was selected using covariate balancing propensity scores [12] with multinomial specification in order to generate the probability of receiving immediate MBSR, Daily Examen, or Stress Proofing interventions, or being designated as a waitlist participant. Propensity score models were generated separately for use in C-SOSI and GAD-7 outcomes vs the PHQ-8 outcome vs the HRV outcomes so that baseline outcome levels could be used in the prediction of treatment receipt (the PHQ-8 outcome is missing in more participants than the C-SOSI and GAD-7 outcomes; the HRV outcomes are available only on a subset of participants). Separate models were also produced for trial vs. observational data. Analyses were performed using an as-treated estimand to reflect the combination of observational with randomized data.

Distributions of propensity scores for participants receiving each of the immediate interventions as well as the waitlist control were visualized using histograms. Covariate balance pre/post propensity score regression adjustment was assessed using regressions with indicators for intervention and covariate adjustment for the propensity score [13].

**Propensity score methods for sensitivity analyses including the observational data**

Propensity score and outcome regression specifications were run for the combined cohort and remained largely the same with the exception of an addition of a binary indicator in the propensity score models to flag participants that were part of the fully observational data vs those that provided trial period data. Post-waitlist baseline and pre-intervention baseline covariate values were combined with the original trial period baseline data to generate the propensity scores, participants that provided waitlist data and intervention period data had two baseline observations that contributed to the computation of the propensity scores.

**Supplemental Methods Table A3**. Baseline variables included in propensity score model, by analytic sample

| **Baseline Variable** | **Trial Phase** | | | **Pooled Trial and Post-Trial Phase** | | |
|---|---|---|---|---|---|---|
| | **C-SOSI and GAD-7** | **PHQ-8** | **HRV** | **C-SOSI and GAD-7** | **PHQ-8** | **HRV** |
| Age (range 26-80) | X | X | X | X | X | X |
| Sex (Male/Female) | X | X | X | X | X | X |
| Single-racial, non-Hispanic White (Yes/No) | X | X | X | X | X | X |
| Married or cohabitating with partner (Yes/No) | X | X | X | X | X | X |
| Any children at home (Yes/No) | X | X | X | X | X | X |
| Full-time clergy in UMC (Yes/No) | X | X | X | X | X | X |
| Bi-vocational (Yes/No) | X | X | X | X | X | X |
| Number of congregations appointed to (None/1/2+) | X | X | X | X | X | X |
| Number of congregants pastored (None/1-149/150+) | X | X | X | X | X | X |
| Hours per week worked as UMC clergy (range 0-80) | X | X | X | X | X | X |
| Alcoholic drink intake (range 0-5) | X | X | X | X | X | X |
| Caffeinated beverage intake (range 0-4) | X | X | X | X | X | X |
| Metabolic equivalents (METs) per week (range 0-476) | X | X | X | X | X | X |
| Practicing Daily Examen 3+ times a week at registration (Yes/No) | X | X | X | X | X | X |
| Practicing Mindfulness 3+ times a week at registration (Yes/No or missing) | X | X | X | X | X | X |
| Number of top preferences among Selah interventions (0/1/2) | X | X | X | X | X | X |
| Body Mass Index (range 18.1-55.8) | X | X | X | X | X | X |
| Self-endorsed high cholesterol (Yes, current or history/Never or missing) | X | X | X | X | X | X |
| Overall life stress at registration (range 0-4) | X | X | X | X | X | X |
| C-SOSI stress symptoms (range 0.04-3.27) | X | | X | X | | X |
| Self-endorsed anxiety (Yes, current or history/Never or missing) | X | X | X | X | X | X |
| GAD-7 anxiety symptoms (range 0-20) | X | | X | X | | X |
| Self-endorsed depression (Yes, current or history/Never or missing) | X | X | X | X | X | X |
| PHQ-8 depression symptoms (range 0-21) | | X | X | | X | X |
| HRV MESOR (range 7.2-128.7) | | | X | | | X |
| HRV Amplitude (range 0.4-70.5) | | | X | | | X |
| Late registrants (Yes/No) | | | | X | X | X |
| In post-trial sample (Yes/No) | | | | X | X | X |

**Model Building for Final Outcome Regression Models**

Invitations for the waitlist surveys (in groups of approximately 20 to be in comparable in size to workshop groups) were agnostic to intervention preference or assignment, which raised concerns about the potential for time confounding. Thus, regressions were also adjusted for number of months from the start of overall survey data collection (April 2020) to each respective survey. We explored the functional form of time from options of linear, square, and cubic, using baseline levels of C-SOSI and GAD-7 scores (to ensure the interventions did not influence time trends), selecting the functional form with the best fit using Akaike Information Criterion (AIC) [14], confirming the plausibility of functional forms using lowess plots.

We additionally explored the possibility that random slopes for study time (weeks from baseline) may better account for within –person correlation over time, however, comparisons of AIC indicated that random intercepts alone performed as well or better than models that included individual level random slopes for study time.

Visualization of residuals was used to assess normality to confirm that the assumptions of the linear model were adequately met.

**Details of Missing Data Methodology**

Our base sample was composed of participants providing any survey or HRV data to the study. Missing data could arise via missing outcome data at any time point (baseline, 12-weeks, or 24-weeks) or missing baseline covariate data (needed for propensity score generation). With missing data greater than 5% on both outcome and covariates and missing completely at random (MCAR) not a plausible (e.g. participants with more stress are plausibly more likely to withdraw from the study) and missing not at random (MNAR) also not a plausible assumption given the richness of available data, multiple imputation using chained equations (MICE) was used [15], [16], [17] to produce an alternative set of estimates as a sensitivity analysis to be compared to the original complete case estimates. The imputation process was integrated in the analytic process using the following steps:

1. Imputation using MICE for all variable included in either the final regression model OR the propensity score model OR that could help predict missing values or the presence of missing data (see Table A2). Perform augmented regression in the presence of perfect prediction for categorical variables. Create 10 imputed datasets
2. Calculate propensity scores separately for each of the 10 imputation datasets as well as the original
3. Perform final regression analyses separately by imputation dataset and combine using Rubin’s rules [18].

In trials with clustered data, ideally the imputation process would incorporate the clustered structure of the data into the imputation process in order to avoid increased risk of Type I error [19]. However, software limitations, multiple treatment groups, and low sample size made incorporation of clustering infeasible. Therefore, the central goal of the imputation process is to ascertain whether there was potential bias in the magnitude of the treatment effect, not to ascertain statistical significance.

**Supplemental Methods Table A4**. Variables included and types of regression used for multiple imputation using chained equations (MICE)

| Variables included | Regression type | C-SOSI and GAD-7 based analytic sample | PHQ-8 based analytic sample | HRV based analytic sample |
|---|---|---|---|---|
| C-SOSI at baseline (range 0.04-3.27) | N/A - independent only | X | | X |
| C-SOSI at 3 months follow-up (range 0-2.58) | Linear | X | | X |
| C-SOSI at 6 months follow-up (range 0-2.44) | Linear | X | | |
| GAD-7 at baseline (range 0-20) | Linear | X | | X |
| GAD-7 at 3 months follow-up (range 0-17) | Linear | X | | X |
| GAD-7 at 6 months follow-up (range 0-15) | Linear | X | | |
| GAD-7 in 2019 from the Panel study (range 0-20) | Linear | X | | |
| GAD-7 in 2021 from the Panel study (range 0-21) | Linear | X | | |
| PHQ-8 at baseline (range 0-21) | Linear | | X | X |
| PHQ-8 at 3 months follow-up (range 0-17) | Linear | | X | X |
| PHQ-8 at 6 months follow-up (range 0-16) | Linear | | X | |
| PHQ-8 in 2019 from the Panel study (range 0-18) | Linear | | X | |
| PHQ-8 in 2021 from the Panel study (range 0-20) | Linear | | X | |
| Metabolic equivalents (METs) per week at baseline (range 0-476) | Linear | X | X | X |
| Hours per week worked as UMC clergy at baseline (range 0-80) | Linear | X | X | X |
| Hours per week worked as UMC clergy in 2019 from the Panel study (range 10-80) | Linear | X | X | |
| Hours per week worked as UMC clergy in 2021 from the Panel study (range 0-80) | Linear | X | X | |
| Number of weeks from baseline to 3 months follow-up (range 7-19) | Linear | X | X | X |
| Number of weeks from baseline to 6 months follow-up (range 8-30) | Linear | X | X | |

| | | | | |
|---|---|---|---|---|
| Number of months from Selah month 1 on calendar to baseline (range 0-12) | N/A - independent only | X | X | X |
| Number of months from Selah month 1 on calendar to 3 months follow-up (range 3-15) | Linear | X | X | X |
| Number of months from Selah month 1 on calendar to 6 months follow-up (range 6-15) | Linear | X | X | |
| Any children at home at baseline (Yes/No) | Logistic | X | X | X |
| Any children at home in 2019 from the Panel study (Yes/No) | Logistic | X | X | |
| Alcoholic drink intake at baseline (range 0-5) | Ordered logistic | X | X | X |
| Caffeinated beverage intake at baseline (range 0-4) | Ordered logistic | X | X | X |
| Number of congregants pastored at baseline (None/1-149/150+) | Ordered logistic | X | X | X |
| Number of congregants pastored in 2019 from the Panel study (range 18-1,400) | Linear | X | X | |
| Number of congregants pastored in 2021 from the Panel study (range 0-1,400) | Linear | X | X | |
| HRV MESOR at baseline (range 7.2-128.7) | Linear | | | X |
| HRV MESOR at 3 months follow-up (range 5.7-105.2) | Linear | | | X |
| HRV amplitude at baseline (range 0.4-70.5) | Linear | | | X |
| HRV amplitude at 3 months follow-up (range 0.4-82.1) | Linear | | | X |
| Overall life stress at registration (range 0-4) | N/A - independent only | X | X | X |
| Self-endorsed depression at baseline (Yes, current or history/Never or missing) | N/A - independent only | X | X | X |
| BMI at baseline (range 18.1-55.8) | N/A - independent only | X | X | X |
| Age at baseline (range 26-80) | N/A - independent only | X | X | X |

| | | | | |
|---|---|---|---|---|
| Female | N/A - independent only | X | X | X |
| As-treated intervention arm | N/A - independent only | X | X | X |

Note: these multiple imputations were done in the Trial sample and not the pooled sample

**References**


1. Tarvainen MP, Niskanen J-P, Lipponen JA, Ranta-Aho PO, Karjalainen PA. Kubios HRV--heart ate variability analysis software. Comput Methods Programs Biomed.
2. Tarvainen M, Lipponen J, Niskanen J, Ranta-aho P. Kubios HRV Software Users Guide [Internet]. 2020 [cited 2021 May 18]. Available from: https://www.kubios.com/downloads/Kubios_HRV_Users_Guide.pdf.
3. Penttilä J, Helminen A, Jartti T, Kuusela T, Huikuri HV, Tulppo MP, et al. Time domain, geometrical and frequency domain analysis of cardiac vagal outflow: effects of various respiratory patterns. Clin Physiol. 2001 May;21(3):365–76.
4. Refinetti R, Lissen GC, Halberg F. Procedures for numerical analysis of circadian rhythms. Biol Rhythm Res [Internet]. 2007 [cited 2021 May 18];38(4):275–325. Available from: https://www.ncbi.nlm.nih.gov/pmc/articles/PMC3663600/
5. Godin G. The Godin-Shephard Leisure-Time Physical Activity Questionnaire. Health Fit J Can [Internet]. 2011 [cited 2021 May 18];4(1):18–22. Available from: https://hfjc.library.ubc.ca/index.php/HFJC/article/view/82
6. National Heart, Lung, and Blood Institute. Clinical guidelines on the identification, evaluation, and treatment of overweight and obesity in adults. Bethesda (MD): National Heart, Lung, and Blood Institute; 1998.
7. Carlson LE, Thomas BC. Development of the Calgary Symptoms of Stress Inventory (C-SOSI). Int J Behav Med. 2007;14(4):249–56.
8. Sidani S, Epstein DR, Bootzin RR, Moritz P, & Miranda, J. (2009). Assessment of preferences for treatment: validation of a measure. Research in nursing & health, 32(4), 419-431
9. https://doi.org/10.1002/sim.6207Laborde S, Mosley, E, & Thayer, JF. (2017). Heart rate variability and cardiac vagal tone in psychophysiological research–recommendations for experiment planning, data analysis, and data reporting. *Frontiers in psychology, 8, 213.*
10. Rubin M. When to adjust alpha during multiple testing: A consideration of disjunction, conjunction, and individual testing. Synthese. 2021
11. Vansteelandt, S, & Daniel, RM. (2014), On regression adjustment for the propensity score, *Statistics in Medicine*, 33, pages 4053– 4072, doi: 10.1002/sim.6207
12. Imai K, Ratkovic M. Covariate balancing propensity score. J Royal Statistical Soc B. 2014;76:243–63.
13. Spreeuwenberg MD, Bartak A, Croon MA, Hagenaars JA, Busschbach JJV, Andrea H, et al. The Multiple propensity score as control for bias in the comparison of more than two treatment arms: An introduction from a case study in mental health on JSTOR. Med Care. 2010;48:166–74.

14. Akaike H. A new look at the statistical model identification. IEEE Trans Automat Contr. 1974;19:716–23.
15. Jakobsen JC, Gluud C, Wetterslev J, Winkel P. When and how should multiple imputation be used for handling missing data in randomised clinical trials - a practical guide with flowcharts. BMC Med Res Methodol. 2017;17:162.
16. van Buuren S. Multiple imputation of discrete and continuous data by fully conditional specification. Stat Methods Med Res. 2007;16(3):219-242. doi:10.1177/0962280206074463
17. Lee KJ, Carlin JB. Multiple imputation for missing data: fully conditional specification versus multivariate normal imputation. Am J Epidemiol. 2010;171(5):624-632. doi:10.1093/aje/kwp425
18. Rubin DB. Multiple Imputation for nonresponse in surveys. Vol 81. Subsequent. Hoboken, N.J: Wiley-interscience; 2004:258.
19. Taljaard M, Donner A, Klar N. Imputation strategies for missing continuous outcomes in cluster randomized trials. Biom J. 2008;50:329–45.

### Appendix B. Supplemental Results

**Supplemental Figure B1**. Flow Chart for Trial Participants During the Trial Period and Post-waitlist Period

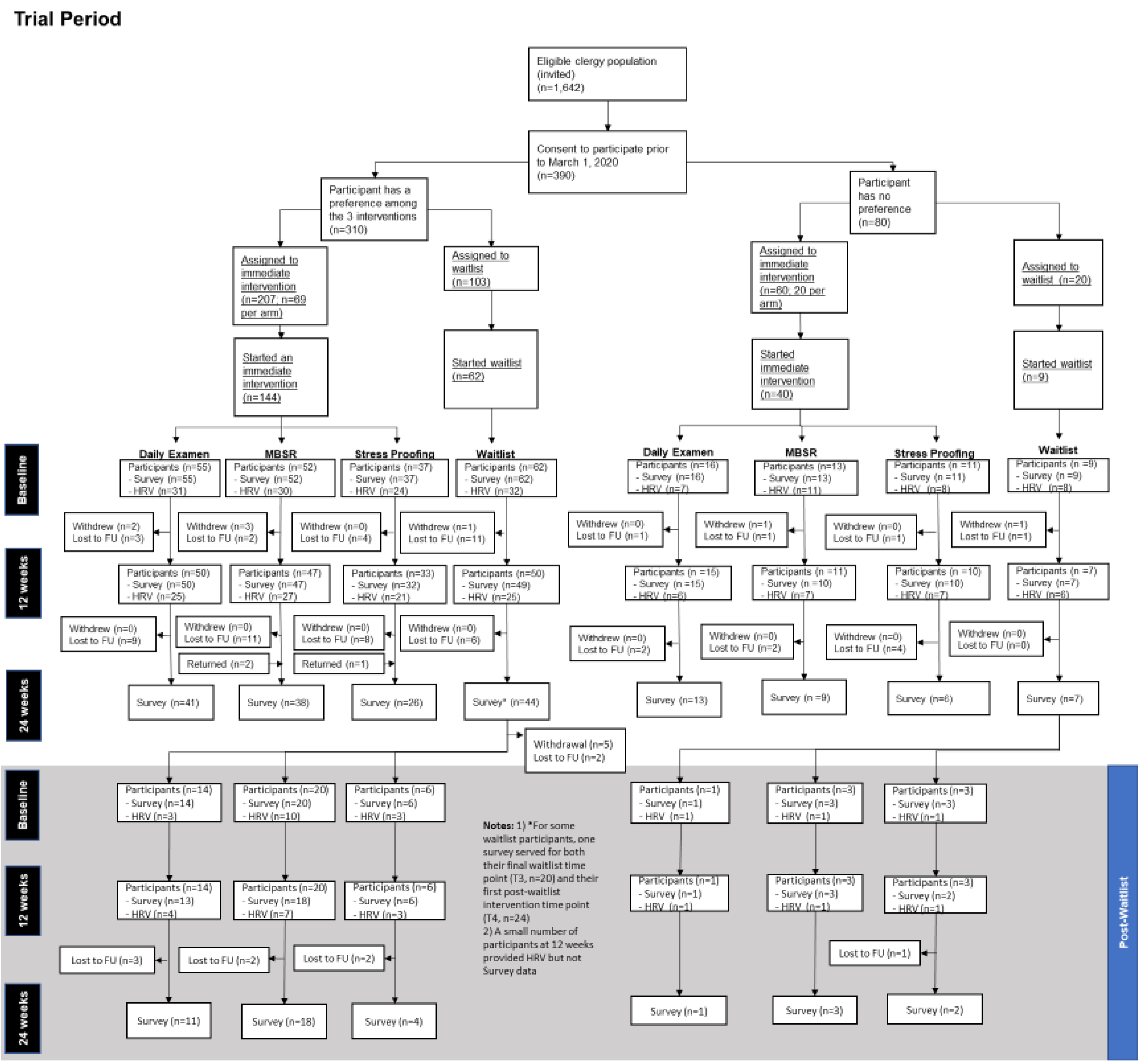


Note: Post-waitlist period data are analyzed as observational data.

**Supplemental Figure B2**. Flow Chart for Observational Participants

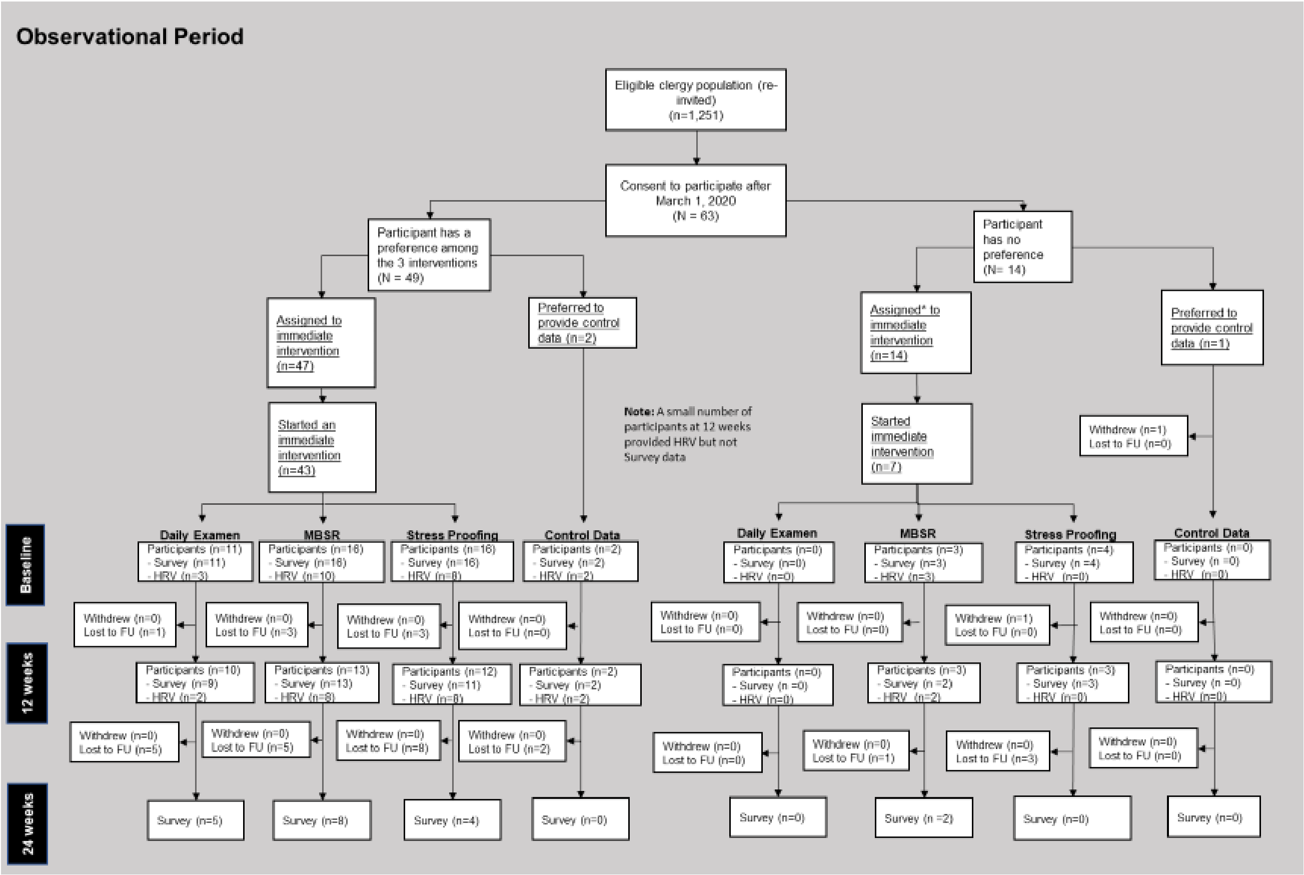

**Supplemental Table B1.** Baseline characteristics of Selah trial phase participants compared to 2019 Clergy Health Panel Study participants[1]

| | Selah total (N = 255) | 2019 Panel Study Population (N = 1278) | *p*-value[2] |
|---|---|---|---|
| **Age (in years)** | | | 0.387 |
| Mean (SD) | 53.9 (11.2) | 53.2 (12.3) | |
| **Sex, n (%)** | | | <0.001 |
| Female | 121 (47.5%) | 429 (33.7%) | |
| Male | 134 (52.5%) | 843 (66.3%) | |
| **Race & ethnicity (in mutual exclusive categories), n (%)** | | | 0.892 |
| White and not Latinx | 231 (90.6%) | 1091 (88.8%) | |
| African American and not Latinx | 15 (5.9%) | 72 (5.9%) | |
| Asian American/Pacific Islander and not Latinx | 2 (0.8%) | 17 (1.4%) | |
| Native American and not Latinx | 2 (0.8%) | 13 (1.1%) | |
| Latinx | 2 (0.8%) | 18 (1.5%) | |
| Other, including bi-/multi-racial | 3 (1.2%) | 18 (1.5%) | |
| **Marital & habitation status, n (%)** | | | 0.183 |
| Not married, or married but separated/divorcing | 27 (10.6%) | 174 (13.7%) | |
| Married or cohabitating | 228 (89.4%) | 1098 (86.3%) | |
| **Any children living at home, n (%)** | | | 0.148 |
| No | 134 (53.2%) | 738 (58.1%) | |
| Yes | 118 (46.8%) | 532 (41.9%) | |
| **Clergy appointment, n (%)** | | | 0.074 |
| Pastoral charge | 210 (82.4%) | 1107 (86.6%) | |
| Extension or other | 45 (17.6%) | 171 (13.4%) | |
| **Bi-vocational, n (%)** | | | <0.001 |
| No | 246 (96.5%) | 1115 (87.6%) | |
| Yes | 9 (3.5%) | 158 (12.4%) | |
| **Hours per week worked as full-time clergy** | | | 0.521 |
| Mean (SD) | 49.5 (10.5) | 49.9 (8.6) | |
| **Financial stress, n (%)** | | | 0.089 |
| Not at all or slightly stressful | 172 (68.8%) | 792 (63.2%) | |
| Moderately, very, or extremely | 78 (31.2%) | 462 (36.8%) | |
| **Body Mass Index (BMI)** | | | 0.088 |
| Mean (SD) | 30.8 (7.0) | 30.0 (6.7) | |
| **Obesity, n (%)** | | | 0.043 |
| Not obese (BMI <30) | 131 (51.4%) | 728 (58.2%) | |
| Obese (BMI 30+) | 124 (48.6%) | 522 (41.8%) | |
| **Hypertension, n (%)** | | | 0.076 |
| No (including missing) | 167 (65.5%) | 761 (59.5%) | |

| | | | |
|---|---|---|---|
| Yes, current or history | 88 (34.5%) | 517 (40.5%) | |
| **Diabetes, n (%)** | | | 0.002 |
| No (including missing) | 223 (87.5%) | 1012 (79.2%) | |
| Yes, current or history | 32 (12.5%) | 266 (20.8%) | |
| **PHQ-8 depression symptoms sum score, [0-24]** | | | <0.001 |
| Mean (SD) | 5.4 (4.6) | 4.2 (4.1) | |
| **Depression screens, n (%)** | | | 0.005 |
| Negative (PHQ8 <10) | 210 (83.0%) | 1133 (89.3%) | |
| Positive (PHQ8 10+) | 43 (17.0%) | 136 (10.7%) | |

[1]Comparison data are from the Clergy Health Longitudinal Survey 2019 wave (73% response rate), of the same study population from which the Selah Study recruited. Comparison participants were appointed and actively serving as clergy at the time of the survey.

[2]*P*-values generated using Kruskal Wallis test for continuous variables chi square text for categorical variables.

**Supplemental Table B2.** Baseline characteristics of immediate intervention and waitlist study arms for trial phase participants, for qualified individuals consenting to give HRV data

| | Waitlist (N = 40) | Stress-Proofing (N = 32) | Daily Examen (N = 38) | Mindfulness-based Stress Reduction (N = 41) | Total (N = 151) |
|---|---|---|---|---|---|
| **Age (in years)** | | | | | |
| Mean (SD) | 53.6 (10.5) | 53.6 (10.5) | 53.8 (12.4) | 52.9 (11.4) | 53.5 (11.2) |
| **Sex, n (%)** | | | | | |
| Female | 18 (45.0%) | 21 (65.6%) | 17 (44.7%) | 20 (48.8%) | 76 (50.3%) |
| Male | 22 (55.0%) | 11 (34.4%) | 21 (55.3%) | 21 (51.2%) | 75 (49.7%) |
| **Race/Ethnicity, n (%)** | | | | | |
| White and not Latinx | 36 (90.0%) | 30 (93.8%) | 36 (94.7%) | 39 (95.1%) | 141 (93.4%) |
| African American and not Latinx | 3 (7.5%) | 2 (6.3%) | 1 (2.6%) | 0 (0.0%) | 6 (4.0%) |
| Asian American/Pacific Islander, Native American, Latinx, bi/multi-racial, and other | 1 (2.5%) | 0 (0.0%) | 1 (2.6%) | 2 (4.9%) | 4 (2.6%) |
| **Marital & habitation status, n (%)** | | | | | |
| Not married, separated or divorced | 6 (15.0%) | 6 (18.8%) | 2 (5.3%) | 2 (4.9%) | 16 (10.6%) |
| Married or cohabitating | 34 (85.0%) | 26 (81.3%) | 36 (94.7%) | 39 (95.1%) | 135 (89.4%) |
| **Any children living at home, n (%)** | | | | | |
| No | 20 (50.0%) | 19 (61.3%) | 18 (47.4%) | 15 (36.6%) | 72 (48.0%) |
| Yes | 20 (50.0%) | 12 (38.7%) | 20 (52.6%) | 26 (63.4%) | 78 (52.0%) |
| **Clergy appointment, n (%)** | | | | | |
| Pastoral charge | 33 (82.5%) | 24 (75.0%) | 33 (86.8%) | 35 (85.4%) | 125 (82.8%) |
| Extension or other | 7 (17.5%) | 8 (25.0%) | 5 (13.2%) | 6 (14.6%) | 26 (17.2%) |
| **Bi-vocational, n (%)** | | | | | |
| No | 38 (95.0%) | 31 (96.9%) | 38 (100.0%) | 39 (95.1%) | 146 (96.7%) |
| Yes | 2 (5.0%) | 1 (3.1%) | 0 (0.0%) | 2 (4.9%) | 5 (3.3%) |
| **Hours per week worked as full-time clergy** | | | | | |
| Mean (SD) | 51.5 (11.8) | 51.2 (10.6) | 47.8 (6.3) | 47.7 (11.0) | 49.5 (10.3) |
| **Stress from congregation(s)/work from Nov 2019 to registration, [0-3]** | | | | | |
| Mean (SD) | 1.8 (0.8) | 2.2 (0.8) | 1.8 (0.7) | 1.8 (0.7) | 1.9 (0.8) |
| **Financial stress, n (%)** | | | | | |
| Not at all or slightly stressful | 31 (77.5%) | 20 (64.5%) | 29 (76.3%) | 25 (62.5%) | 105 (70.5%) |
| Moderately, very, or extremely | 9 (22.5%) | 11 (35.5%) | 9 (23.7%) | 15 (37.5%) | 44 (29.5%) |
| **Alcoholic drink intake, n (%)** | | | | | |
| None | 12 (30.0%) | 12 (38.7%) | 9 (24.3%) | 12 (30.8%) | 45 (30.6%) |
| Occasional drink (not every week) | 12 (30.0%) | 6 (19.4%) | 11 (29.7%) | 11 (28.2%) | 40 (27.2%) |
| 1-2 drinks | 7 (17.5%) | 6 (19.4%) | 8 (21.6%) | 8 (20.5%) | 29 (19.7%) |
| 3-6 drinks | 6 (15.0%) | 5 (16.1%) | 4 (10.8%) | 5 (12.8%) | 20 (13.6%) |
| about a drink a day | 2 (5.0%) | 2 (6.5%) | 3 (8.1%) | 3 (7.7%) | 10 (6.8%) |
| more than a drink a day | 1 (2.5%) | 0 (0.0%) | 2 (5.4%) | 0 (0.0%) | 3 (2.0%) |

| | | | | | |
|---|---|---|---|---|---|
| **Self-reported current heavy alcohol use, n (%)** | | | | | |
| No | 39 (100.0%) | 30 (96.8%) | 35 (94.6%) | 38 (100.0%) | 142 (97.9%) |
| Yes | 0 (0.0%) | 1 (3.2%) | 2 (5.4%) | 0 (0.0%) | 3 (2.1%) |
| **Caffeinated beverage intake per day, n (%)** | | | | | |
| None | 4 (10.0%) | 5 (16.1%) | 3 (8.1%) | 5 (12.8%) | 17 (11.6%) |
| 1 cup | 11 (27.5%) | 5 (16.1%) | 9 (24.3%) | 10 (25.6%) | 35 (23.8%) |
| 2-3 cups | 19 (47.5%) | 14 (45.2%) | 18 (48.6%) | 19 (48.7%) | 70 (47.6%) |
| 4-5 cups | 4 (10.0%) | 6 (19.4%) | 5 (13.5%) | 5 (12.8%) | 20 (13.6%) |
| 6 or more cups | 2 (5.0%) | 1 (3.2%) | 2 (5.4%) | 0 (0.0%) | 5 (3.4%) |
| **Metabolic equivalents (METs) per week** | | | | | |
| Mean (SD) | 63.0 (68.8) | 78.6 (74.9) | 71.8 (89.5) | 46.7 (71.8) | 64.2 (76.7) |
| **Body Mass Index (BMI)** | | | | | |
| Mean (SD) | 30.4 (6.6) | 31.2 (7.7) | 30.0 (6.4) | 30.8 (7.9) | 30.6 (7.1) |
| **Obesity, n (%)** | | | | | |
| Not obese (BMI <30) | 21 (52.5%) | 17 (53.1%) | 20 (52.6%) | 21 (51.2%) | 79 (52.3%) |
| Obese (BMI 30+) | 19 (47.5%) | 15 (46.9%) | 18 (47.4%) | 20 (48.8%) | 72 (47.7%) |
| **High blood pressure, n (%)** | | | | | |
| No (including missing) | 24 (60.0%) | 23 (71.9%) | 23 (60.5%) | 31 (75.6%) | 101 (66.9%) |
| Yes, current or history | 16 (40.0%) | 9 (28.1%) | 15 (39.5%) | 10 (24.4%) | 50 (33.1%) |
| **Diabetes, n (%)** | | | | | |
| No (including missing) | 33 (82.5%) | 29 (90.6%) | 33 (86.8%) | 38 (92.7%) | 133 (88.1%) |
| Yes, current or history | 7 (17.5%) | 3 (9.4%) | 5 (13.2%) | 3 (7.3%) | 18 (11.9%) |
| **PHQ-8 depression symptoms sum score, [0-24]** | | | | | |
| Mean (SD) | 4.3 (4.3) | 5.5 (4.3) | 5.5 (4.5) | 5.7 (4.8) | 5.2 (4.5) |
| **Depression screens, n (%)** | | | | | |
| Negative (PHQ-8 <10) | 36 (90.0%) | 27 (84.4%) | 32 (84.2%) | 35 (85.4%) | 130 (86.1%) |
| Positive (PHQ-8 10+) | 4 (10.0%) | 5 (15.6%) | 6 (15.8%) | 6 (14.6%) | 21 (13.9%) |

**Supplemental Table B3.** Baseline characteristics of pooled trial and observational participants

| | Waitlist | Stress-Proofing | Daily Examen | Mindfulness-based Stress Reduction | Total |
|---|---|---|---|---|---|
| | (N = 73) | (N = 77) | (N = 97) | (N = 107) | (N = 354) |
| **Age (in years)** | | | | | |
| Mean (SD) | 54.5 (10.3) | 52.2 (11.7) | 55.4 (11.4) | 52.0 (11.9) | 53.5 (11.4) |
| **Sex, n (%)** | | | | | |
| Female | 32 (43.8%) | 48 (62.3%) | 41 (42.3%) | 56 (52.3%) | 177 (50.0%) |
| Male | 41 (56.2%) | 29 (37.7%) | 56 (57.7%) | 51 (47.7%) | 177 (50.0%) |
| **Race/Ethnicity, n (%)** | | | | | |
| White and not Latinx | 67 (91.8%) | 64 (83.1%) | 84 (86.6%) | 100 (93.5%) | 315 (89.0%) |
| African American and not Latinx | 5 (6.8%) | 7 (9.1%) | 6 (6.2%) | 4 (3.7%) | 22 (6.2%) |
| Asian-American/Pacific Islander, Native American, Latinx, multi-racial, and other | 1 (1.4%) | 6 (7.8%) | 7 (7.2%) | 3 (2.8%) | 17 (4.8%) |
| **Marital & habitation status, n (%)** | | | | | |
| Not married, or married but separated/divorcing | 8 (11.0%) | 16 (20.8%) | 9 (9.3%) | 12 (11.2%) | 45 (12.7%) |
| Married or cohabitating | 65 (89.0%) | 61 (79.2%) | 88 (90.7%) | 95 (88.8%) | 309 (87.3%) |
| **Any children living at home, n (%)** | | | | | |
| No | 39 (53.4%) | 43 (58.1%) | 59 (61.5%) | 53 (50.5%) | 194 (55.7%) |
| Yes | 34 (46.6%) | 31 (41.9%) | 37 (38.5%) | 52 (49.5%) | 154 (44.3%) |
| **Clergy appointment, n (%)** | | | | | |
| Pastoral charge | 59 (80.8%) | 61 (79.2%) | 82 (84.5%) | 85 (79.4%) | 287 (81.1%) |
| Extension or other | 14 (19.2%) | 16 (20.8%) | 15 (15.5%) | 22 (20.6%) | 67 (18.9%) |
| **Bi-vocational, n (%)** | | | | | |
| No | 70 (95.9%) | 71 (93.4%) | 93 (95.9%) | 104 (97.2%) | 338 (95.8%) |
| Yes | 3 (4.1%) | 5 (6.6%) | 4 (4.1%) | 3 (2.8%) | 15 (4.2%) |
| **Hours per week worked as full-time clergy** | | | | | |
| Mean (SD) | 49.4 (9.9) | 48.7 (10.8) | 49.5 (10.6) | 49.0 (10.2) | 49.2 (10.3) |
| **Stress from congregation(s)/work from Nov 2019 to registration, [0-3]** | | | | | |
| Mean (SD) | 1.8 (0.7) | 2.0 (0.8) | 1.8 (0.6) | 1.9 (0.7) | 1.9 (0.7) |
| **Financial stress, n (%)** | | | | | |
| Not at all or slightly stressful | 50 (69.4%) | 48 (64.9%) | 68 (70.1%) | 71 (68.3%) | 237 (68.3%) |
| Moderately, very, or extremely | 22 (30.6%) | 26 (35.1%) | 29 (29.9%) | 33 (31.7%) | 110 (31.7%) |
| **Alcoholic drink intake, n (%)** | | | | | |
| None | 23 (31.9%) | 21 (28.4%) | 36 (37.5%) | 33 (32.0%) | 113 (32.8%) |
| Occasional drink (not every week) | 17 (23.6%) | 25 (33.8%) | 23 (24.0%) | 28 (27.2%) | 93 (27.0%) |
| 1-2 drinks | 11 (15.3%) | 11 (14.9%) | 17 (17.7%) | 17 (16.5%) | 56 (16.2%) |
| 3-6 drinks | 13 (18.1%) | 11 (14.9%) | 8 (8.3%) | 16 (15.5%) | 48 (13.9%) |
| about a drink a day | 6 (8.3%) | 4 (5.4%) | 8 (8.3%) | 8 (7.8%) | 26 (7.5%) |
| more than a drink a day | 2 (2.8%) | 2 (2.7%) | 4 (4.2%) | 1 (1.0%) | 9 (2.6%) |

| | | | | | |
|---|---|---|---|---|---|
| **Self-reported current heavy alcohol use, n (%)** | | | | | |
| No | 70 (100.0%) | 71 (97.3%) | 93 (96.9%) | 101 (99.0%) | 335 (98.2%) |
| Yes | 0 (0.0%) | 2 (2.7%) | 3 (3.1%) | 1 (1.0%) | 6 (1.8%) |
| **Caffeinated beverage intake per day, n (%)** | | | | | |
| None | 9 (12.5%) | 7 (9.5%) | 12 (12.5%) | 11 (10.7%) | 39 (11.3%) |
| 1 cup | 18 (25.0%) | 21 (28.4%) | 16 (16.7%) | 30 (29.1%) | 85 (24.6%) |
| 2-3 cups | 33 (45.8%) | 33 (44.6%) | 54 (56.3%) | 46 (44.7%) | 166 (48.1%) |
| 4-5 cups | 10 (13.9%) | 12 (16.2%) | 10 (10.4%) | 12 (11.7%) | 44 (12.8%) |
| 6 or more cups | 2 (2.8%) | 1 (1.4%) | 4 (4.2%) | 4 (3.9%) | 11 (3.2%) |
| **Metabolic equivalents (METs) per week** | | | | | |
| Mean (SD) | 70.0 (89.1) | 53.6 (64.6) | 80.1 (99.7) | 50.7 (68.1) | 63.4 (82.3) |
| **Body Mass Index (BMI)** | | | | | |
| Mean (SD) | 30.4 (6.9) | 31.0 (6.7) | 30.4 (6.5) | 30.9 (7.4) | 30.7 (6.9) |
| **Obesity, n (%)** | | | | | |
| Not obese (BMI <30) | 39 (53.4%) | 34 (44.2%) | 49 (50.5%) | 55 (51.4%) | 177 (50.0%) |
| Obese (BMI 30+) | 34 (46.6%) | 43 (55.8%) | 48 (49.5%) | 52 (48.6%) | 177 (50.0%) |
| **High blood pressure, n (%)** | | | | | |
| No (including missing) | 45 (61.6%) | 52 (67.5%) | 63 (64.9%) | 77 (72.0%) | 237 (66.9%) |
| Yes, current or history | 28 (38.4%) | 25 (32.5%) | 34 (35.1%) | 30 (28.0%) | 117 (33.1%) |
| **Diabetes, n (%)** | | | | | |
| No (including missing) | 60 (82.2%) | 72 (93.5%) | 81 (83.5%) | 99 (92.5%) | 312 (88.1%) |
| Yes, current or history | 13 (17.8%) | 5 (6.5%) | 16 (16.5%) | 8 (7.5%) | 42 (11.9%) |
| **PHQ-8 depression symptoms sum score, [0-24]** | | | | | |
| Mean (SD) | 4.2 (3.9) | 5.8 (4.6) | 5.1 (4.3) | 6.4 (5.2) | 5.4 (4.6) |
| **Depression screens, n (%)** | | | | | |
| Negative (PHQ-8 <10) | 64 (88.9%) | 59 (76.6%) | 83 (85.6%) | 80 (75.5%) | 286 (81.3%) |
| Positive (PHQ-8 10+) | 8 (11.1%) | 18 (23.4%) | 14 (14.4%) | 26 (24.5%) | 66 (18.8%) |

Note. Of the sample of 354, some participants had dual baselines; there were 307 total unique participants (73 were initially assigned to the waitlist, 68 to Stress Proofing, 82 to Daily Examen, and 84 to Mindfulness-based Stress Reduction)

**Supplemental Table B4**. Intervention attendance and participation

| | Trial participants (N = 184) | Pooled Trial and Observational Participants (N = 234) |
|---|---|---|
| **Number of Stress Proofing main sessions attended (Range 0-4)** | | |
| 0 | 0 (0.0%) | 2 (2.9%) |
| 1 | 2 (4.2%) | 3 (4.4%) |
| 2 | 4 (8.3%) | 6 (8.8%) |
| 3 | 12 (25.0%) | 14 (20.6%) |
| 4 | 30 (62.5%) | 43 (63.2%) |
| **Did the participant attend the Stress Proofing follow-up session?** | | |
| No | 18 (42.9%) | 27 (46.6%) |
| Yes | 24 (57.1%) | 31 (53.4%) |
| **Number of Daily Examen main sessions attended (Range 0-3)** | | |
| 0 | 0 (0.0%) | 0 (0.0%) |
| 1 | 0 (0.0%) | 0 (0.0%) |
| 2 | 3 (4.2%) | 4 (4.9%) |
| 3 | 68 (95.8%) | 78 (95.1%) |
| **Number of Daily Examen follow-up sessions attended (Range 0-2)** | | |
| 0 | 34 (47.9%) | 39 (47.6%) |
| 1 | 18 (25.4%) | 23 (28.0%) |
| 2 | 19 (26.8%) | 20 (24.4%) |
| **Number of Mindfulness-based Stress Reduction main sessions attended (Range 0-8)** | | |
| Mean (SD) | 6.9 (1.5) | 7.0 (1.4) |
| Median (Q1, Q3) | 7.0 (7.0, 8.0) | 7.0 (7.0, 8.0) |

**Supplemental Figure B3.** Text message response rates, practice responses, and trends over 24 weeks

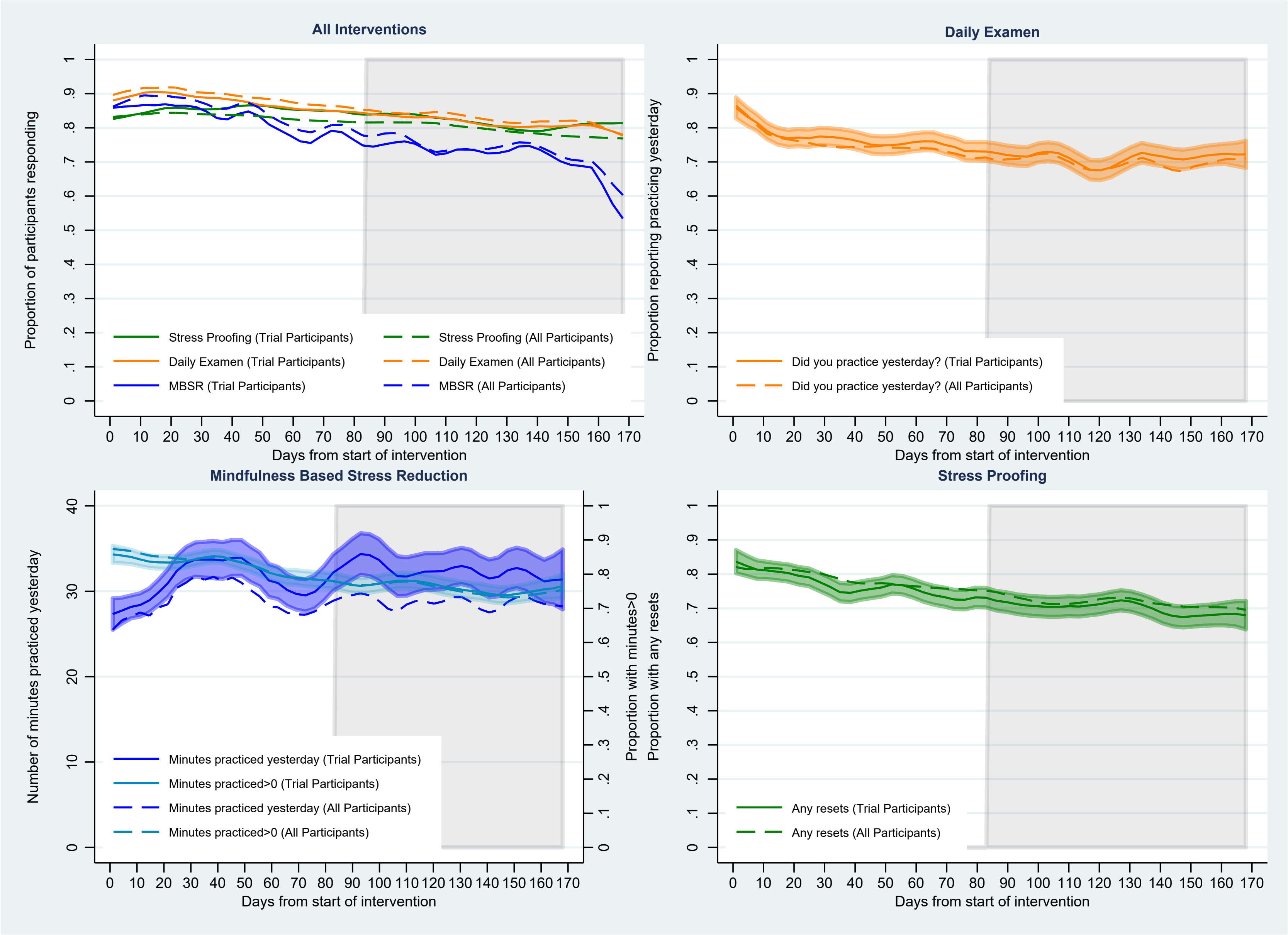

**Supplemental Figure B4.** Distribution of propensity scores by treatment condition, outcome, and population type

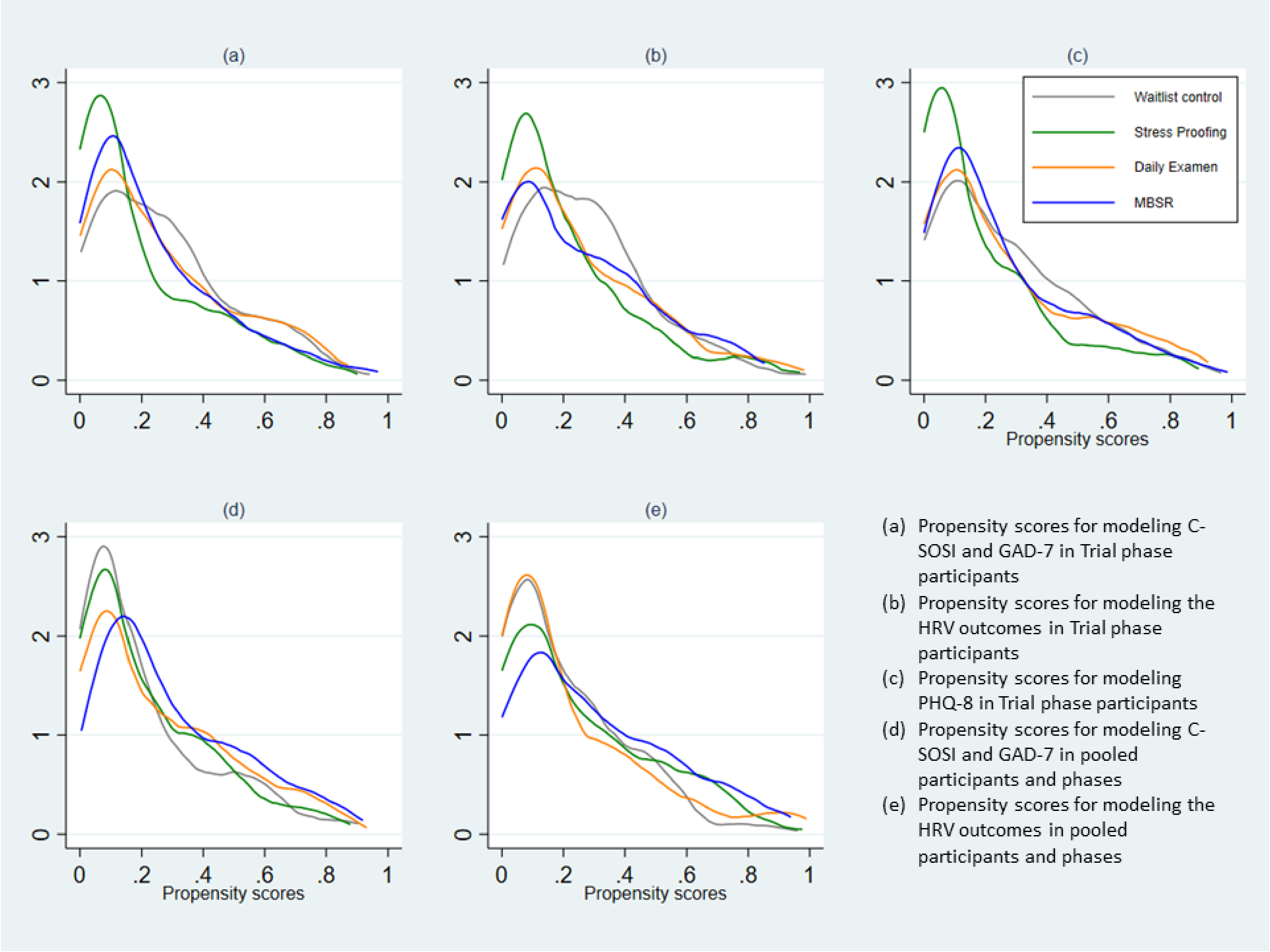

**Supplemental Table B5.** Baseline characteristic balance between treatment conditions before and after propensity score adjustment for C-SOSI and anxiety outcomes in trial participants

| | Unadjusted Mean (SD) or % (n) | | | | *P-value* | |
|---|---|---|---|---|---|---|
| | Stress Proofing | Daily Examen | Mindfulness-based Stress Reduction | Waitlist | Before adjustment | After adjustment |
| Age (in years), mean (SD) | 53.2 (10.7) | 54.0 (12.1) | 52.7 (11.7) | 54.7 (10.1) | .784 | .854 |
| Sex, n (%) | | | | | .169 | .335 |
| Male | 37.0% (17) | 58.7% (37) | 51.7% (30) | 51.5% (34) | | |
| Female | 63.0% (29) | 41.3% (26) | 48.3% (28) | 48.5% (32) | | |
| Race & ethnicity, n (%) | | | | | .604 | .826 |
| Single-racial, non-Latinx white | 93.5% (43) | 87.3% (55) | 93.1% (54) | 92.4% (61) | | |
| All other ethnicities, including bi-/multi-racial | 6.5% (3) | 12.7% (8) | 6.9% (4) | 7.6% (5) | | |
| Marital & habitation status, n (%) | | | | | .157 | .366 |
| Not married, or married but separated/divorcing | 17.4% (8) | 6.3% (4) | 5.2% (3) | 12.1% (8) | | |
| Married or cohabitating with partner | 82.6% (38) | 93.7% (59) | 94.8% (55) | 87.9% (58) | | |
| Any children living at home, n (%) | | | | | .374 | .705 |
| No | 58.7% (27) | 54.0% (34) | 43.1% (25) | 56.1% (37) | | |
| Yes | 41.3% (19) | 46.0% (29) | 56.9% (33) | 43.9% (29) | | |
| Full-time clergy, n (%) | | | | | .460 | .745 |
| No | 15.2% (7) | 20.6% (13) | 10.3% (6) | 13.6% (9) | | |
| Yes | 84.8% (39) | 79.4% (50) | 89.7% (52) | 86.4% (57) | | |
| Bi-vocational, n (%) | | | | | .816 | .991 |
| No | 95.7% (44) | 98.4% (62) | 96.6% (56) | 95.5% (63) | | |
| Yes | 4.3% (2) | 1.6% (1) | 3.4% (2) | 4.5% (3) | | |
| Number of congregations appointed to, n (%) | | | | | .589 | .704 |
| Not appointed to a local congregation | 17.4% (8) | 12.7% (8) | 15.5% (9) | 16.7% (11) | | |
| 1 congregation | 52.2% (24) | 65.1% (41) | 63.8% (37) | 68.2% (45) | | |
| 2+ congregations | 30.4% (14) | 22.2% (14) | 20.7% (12) | 15.2% (10) | | |
| Number of congregants pastored, n (%) | | | | | .501 | .795 |
| Not appointed to a local congregation | 19.6% (9) | 15.9% (10) | 20.7% (12) | 18.2% (12) | | |

| | | | | | | |
|---|---|---|---|---|---|---|
| 1-149 people in worship per week | 60.9% (28) | 68.3% (43) | 60.3% (35) | 51.5% (34) | | |
| 150+ people in worship per week | 19.6% (9) | 15.9% (10) | 19.0% (11) | 30.3% (20) | | |
| Hours per week worked as clergy, mean (SD) | 46.5 (13.8) | 44.7 (13.6) | 47.1 (13.5) | 47.2 (11.8) | .686 | .747 |
| Alcoholic drink intake, scale range 0-5, mean (SD) | 1.35 (1.35) | 1.24 (1.34) | 1.47 (1.39) | 1.50 (1.46) | .711 | .900 |
| Caffeinated beverage intake, scale range 0-4, mean (SD) | 1.78 (0.89) | 1.67 (0.95) | 1.76 (1.00) | 1.70 (0.96) | .913 | .990 |
| Metabolic equivalents (METs) per week, mean (SD) | 58.4 (62.6) | 74.6 (85.7) | 58.4 (77.8) | 70.8 (91.8) | .609 | .979 |
| Practicing Daily Examen 3+ times a week at registration, n (%) | | | | | .883 | .937 |
| No | 93.5% (43) | 90.5% (57) | 93.1% (54) | 93.9% (62) | | |
| Yes | 6.5% (3) | 9.5% (6) | 6.9% (4) | 6.1% (4) | | |
| Practicing Mindfulness 3+ times a week at registration, n (%) | | | | | .532 | .711 |
| No or missing | 93.5% (43) | 90.5% (57) | 96.6% (56) | 95.5% (63) | | |
| Yes | 6.5% (3) | 9.5% (6) | 3.4% (2) | 4.5% (3) | | |
| Number of top preferences among Selah interventions, n (%) | | | | | .296 | .817 |
| No preference | 21.7% (10) | 25.4% (16) | 20.7% (12) | 12.1% (8) | | |
| 1 top preference | 76.1% (35) | 68.3% (43) | 69.0% (40) | 81.8% (54) | | |
| 2 tied preferences | 2.2% (1) | 6.3% (4) | 10.3% (6) | 6.1% (4) | | |
| Body Mass Index, mean (SD) | 31.0 (7.0) | 29.9 (6.4) | 32.0 (8.0) | 30.4 (7.1) | .431 | .987 |
| Self-endorsed high cholesterol, n (%) | | | | | .188 | .823 |
| Never or missing | 73.9% (34) | 54.0% (34) | 56.9% (33) | 59.1% (39) | | |
| Yes, current or history | 26.1% (12) | 46.0% (29) | 43.1% (25) | 40.9% (27) | | |
| Overall life stress at registration, scale range 0-4, mean (SD) | 2.46 (0.75) | 2.32 (0.78) | 2.45 (0.82) | 2.42 (0.91) | .781 | .972 |
| C-SOSI stress symptoms, scale range 0-4, mean (SD) | 1.12 (0.60) | 0.93 (0.56) | 1.16 (0.54) | 0.87 (0.52) | .010 | .498 |
| Self-endorsed single-item anxiety, n (%) | | | | | .016 | .538 |
| Never or missing | 63.0% (29) | 77.8% (49) | 51.7% (30) | 72.7% (48) | | |
| Yes, current or history | 37.0% (17) | 22.2% (14) | 48.3% (28) | 27.3% (18) | | |
| GAD-7 anxiety symptoms, scale range 0-21, mean (SD) | 5.0 (4.0) | 4.6 (4.9) | 6.0 (5.1) | 4.2 (3.9) | .133 | .713 |
| Self-endorsed single-item depression, n (%) | | | | | .015 | .580 |
| Never or missing | 60.9% (28) | 73.0% (46) | 46.6% (27) | 69.7% (46) | | |
| Yes, current or history | 39.1% (18) | 27.0% (17) | 53.4% (31) | 30.3% (20) | | |

**Supplemental Table B6.** Baseline characteristic balance between treatment conditions before and after propensity score adjustment for HRV outcomes in trial participants

| | Unadjusted Mean (SD) or % (n) | | | | *P-value* | |
|---|---|---|---|---|---|---|
| | Stress Proofing | Daily Examen | Mindfulness-Based Stress Reduction | Waitlist | Before adjustment | After adjustment |
| Age (in years), mean (SD) | 53.2 (10.7) | 53.5 (12.6) | 53.3 (10.8) | 53.4 (10.5) | 1.000 | .950 |
| Sex, n (%) | | | | | .281 | .781 |
| Male | 33.3% (10) | 55.6% (20) | 48.6% (18) | 53.8% (21) | | |
| Female | 66.7% (20) | 44.4% (16) | 51.4% (19) | 46.2% (18) | | |
| Race & ethnicity, n (%) | | | | | .832 | .994 |
| Single-racial, non-Hispanic White | 93.3% (28) | 94.4% (34) | 94.6% (35) | 89.7% (35) | | |
| All other ethnicities, including bi-/multi-racial | 6.7% (2) | 5.6% (2) | 5.4% (2) | 10.3% (4) | | |
| Marital & habitation status, n (%) | | | | | .118 | .960 |
| Not married, or married but separated/divorcing | 20.0% (6) | 5.6% (2) | 2.7% (1) | 15.4% (6) | | |
| Married or cohabitating with partner | 80.0% (24) | 94.4% (34) | 97.3% (36) | 84.6% (33) | | |
| Any children at home, n (%) | | | | | .101 | .856 |
| No | 60.0% (18) | 47.2% (17) | 29.7% (11) | 48.7% (19) | | |
| Yes | 40.0% (12) | 52.8% (19) | 70.3% (26) | 51.3% (20) | | |
| Full-time clergy in UMC, n (%) | | | | | .810 | .987 |
| No | 16.7% (5) | 22.2% (8) | 13.5% (5) | 17.9% (7) | | |
| Yes | 83.3% (25) | 77.8% (28) | 86.5% (32) | 82.1% (32) | | |
| Number of congregations appointed to, n (%) | | | | | .419 | .968 |
| Not appointed to a local congregation | 23.3% (7) | 11.1% (4) | 10.8% (4) | 15.4% (6) | | |
| 1 congregation | 43.3% (13) | 69.4% (25) | 67.6% (25) | 64.1% (25) | | |
| 2+ congregations | 33.3% (10) | 19.4% (7) | 21.6% (8) | 20.5% (8) | | |
| Number of congregants pastored, n (%) | | | | | .220 | .970 |
| Not appointed to a local congregation | 26.7% (8) | 13.9% (5) | 16.2% (6) | 17.9% (7) | | |
| 1-149 people in worship per week | 60.0% (18) | 69.4% (25) | 64.9% (24) | 46.2% (18) | | |
| 150+ people in worship per week | 13.3% (4) | 16.7% (6) | 18.9% (7) | 35.9% (14) | | |
| Hours per week worked as UMC clergy, mean (SD) | 47.6 (14.4) | 42.6 (11.7) | 45.3 (13.2) | 47.7 (14.3) | .328 | .967 |

| | | | | | | |
|---|---|---|---|---|---|---|
| Alcoholic drink intake, scale range 0-5, mean (SD) | 1.30 (1.34) | 1.61 (1.46) | 1.35 (1.27) | 1.38 (1.33) | .783 | .850 |
| Caffeinated beverage intake, scale range 0-4, mean (SD) | 1.83 (1.02) | 1.83 (0.97) | 1.59 (0.90) | 1.72 (0.97) | .687 | .957 |
| Metabolic equivalents (METs) per week, mean (SD) | 74.3 (65.2) | 75.8 (90.3) | 47.4 (73.1) | 61.2 (68.7) | .352 | .952 |
| Practicing Daily Examen 3+ times a week at registration, n (%) | | | | | | |
| No | 96.7% (29) | 94.4% (34) | 97.3% (36) | 94.9% (37) | | |
| Yes | 3.3% (1) | 5.6% (2) | 2.7% (1) | 5.1% (2) | | |
| Practicing Mindfulness 3+ times a week at registration, n (%) | | | | | .333 | .942 |
| No or missing | 96.7% (29) | 88.9% (32) | 97.3% (36) | 97.4% (38) | | |
| Yes | 3.3% (1) | 11.1% (4) | 2.7% (1) | 2.6% (1) | | |
| Number of top preferences among Selah interventions, n (%) | | | | | .768 | 1.000 |
| No preference | 23.3% (7) | 19.4% (7) | 27.0% (10) | 17.9% (7) | | |
| 1 top preference | 73.3% (22) | 72.2% (26) | 64.9% (24) | 79.5% (31) | | |
| 2 tied preferences | 3.3% (1) | 8.3% (3) | 8.1% (3) | 2.6% (1) | .768 | 1.000 |
| Body Mass Index, mean (SD) | 31.2 (8.0) | 29.6 (6.1) | 31.3 (8.0) | 30.5 (6.6) | .714 | .985 |
| Self-endorsed high cholesterol, n (%) | | | | | | |
| Never or missing | 76.7% (23) | 58.3% (21) | 59.5% (22) | 56.4% (22) | | |
| Yes, current or history | 23.3% (7) | 41.7% (15) | 40.5% (15) | 43.6% (17) | | |
| Overall life stress at registration, scale range 0-4, mean (SD) | 2.50 (0.78) | 2.39 (0.84) | 2.35 (0.86) | 2.44 (1.05) | .916 | .983 |
| C-SOSI stress symptoms, scale range 0-4, mean (SD) | 1.14 (0.63) | 0.98 (0.55) | 1.07 (0.44) | 0.87 (0.57) | .199 | .863 |
| Self-endorsed anxiety, n (%) | | | | | .118 | .893 |
| Never or missing | 60.0% (18) | 72.2% (26) | 48.6% (18) | 71.8% (28) | | |
| Yes, current or history | 40.0% (12) | 27.8% (10) | 51.4% (19) | 28.2% (11) | | |
| GAD7 anxiety symptoms, scale range 0-21, mean (SD) | 5.0 (3.8) | 4.7 (4.7) | 5.2 (4.7) | 4.2 (4.0) | .782 | .981 |
| Self-endorsed depression, n (%) | | | | | .345 | .998 |
| Never or missing | 60.0% (18) | 66.7% (24) | 48.6% (18) | 66.7% (26) | | |
| Yes, current or history | 40.0% (12) | 33.3% (12) | 51.4% (19) | 33.3% (13) | | |

**Supplemental Table B7.** Baseline characteristic balance between treatment conditions before and after p-value adjustment for C-SOSI and anxiety outcomes in pooled trial and observational participants

| | Unadjusted Mean (SD) or % (n) | | | | *P-value* | |
|---|---|---|---|---|---|---|
| | Stress Proofing | Daily Examen | Mindfulness Based Stress Reduction | Waitlist | Before adjustment | After adjustment |
| Age (in years), mean (SD) | 51.2 (11.3) | 55.6 (11.9) | 52.5 (11.1) | 54.7 (10.1) | .030 | .417 |
| Sex, n (%) | | | | | .055 | .245 |
| Male | 37.2% (29) | 57.0% (57) | 43.8% (49) | 49.1% (52) | | |
| Female | 62.8% (49) | 43.0% (43) | 56.3% (63) | 50.9% (54) | | |
| Race & ethnicity, n (%) | | | | | .227 | .421 |
| Single-racial, non-Latinx white | 87.2% (68) | 88.0% (88) | 94.6% (106) | 92.5% (98) | | |
| All other ethnicities, including bi-/multi-racial | 12.8% (10) | 12.0% (12) | 5.4% (6) | 7.5% (8) | | |
| Marital & habitation status, n (%) | | | | | .318 | .485 |
| Not married, or married but separated/divorcing | 17.9% (14) | 9.0% (9) | 10.7% (12) | 12.3% (13) | | |
| Married or cohabitating with partner | 82.1% (64) | 91.0% (91) | 89.3% (100) | 87.7% (93) | | |
| Any children living at home, n (%) | | | | | .131 | .473 |
| No | 59.0% (46) | 63.0% (63) | 47.3% (53) | 55.7% (59) | | |
| Yes | 41.0% (32) | 37.0% (37) | 52.7% (59) | 44.3% (47) | | |
| Full-time clergy, n (%) | | | | | .201 | .390 |
| No | 15.4% (12) | 21.0% (21) | 10.7% (12) | 13.2% (14) | | |
| Yes | 84.6% (66) | 79.0% (79) | 89.3% (100) | 86.8% (92) | | |
| Bi-vocational, n (%) | | | | | .800 | .792 |
| No | 94.9% (74) | 95.0% (95) | 97.3% (109) | 96.2% (102) | | |
| Yes | 5.1% (4) | 5.0% (5) | 2.7% (3) | 3.8% (4) | | |
| Number of congregations appointed to, n (%) | | | | | .556 | .644 |
| Not appointed to a local congregation | 16.7% (13) | 12.0% (12) | 20.5% (23) | 16.0% (17) | | |
| 1 congregation | 65.4% (51) | 69.0% (69) | 62.5% (70) | 71.7% (76) | | |
| 2+ congregations | 17.9% (14) | 19.0% (19) | 17.0% (19) | 12.3% (13) | | |
| Number of congregants pastored, n (%) | | | | | .243 | .403 |
| Not appointed to a local congregation | 20.5% (16) | 15.0% (15) | 23.2% (26) | 17.9% (19) | | |

| | | | | | | |
|---|---|---|---|---|---|---|
| 1-149 people in worship per week | 55.1% (43) | 67.0% (67) | 56.3% (63) | 51.9% (55) | | |
| 150+ people in worship per week | 24.4% (19) | 18.0% (18) | 20.5% (23) | 30.2% (32) | | |
| Hours per week worked as clergy, mean (SD) | 45.0 (14.1) | 44.4 (13.3) | 46.9 (12.8) | 47.2 (11.8) | .334 | .475 |
| Alcoholic drink intake, scale range 0-5, mean (SD) | 1.49 (1.37) | 1.21 (1.28) | 1.45 (1.37) | 1.54 (1.47) | .344 | .509 |
| Caffeinated beverage intake, scale range 0-4, mean (SD) | 1.71 (0.88) | 1.78 (0.96) | 1.65 (0.97) | 1.70 (0.97) | .806 | .762 |
| Metabolic equivalents (METs) per week, mean (SD) | 55.6 (59.7) | 85.7 (105.5) | 46.1 (65.0) | 73.3 (97.2) | .004 | .213 |
| Practicing Daily Examen 3+ times a week at registration, n (%) | | | | | .579 | .648 |
| No | 91.0% (71) | 92.0% (92) | 95.5% (107) | 94.3% (100) | | |
| Yes | 9.0% (7) | 8.0% (8) | 4.5% (5) | 5.7% (6) | | |
| Practicing Mindfulness 3+ times a week at registration, n (%) | | | | | .468 | .603 |
| No or missing | 92.3% (72) | 92.0% (92) | 96.4% (108) | 95.3% (101) | | |
| Yes | 7.7% (6) | 8.0% (8) | 3.6% (4) | 4.7% (5) | | |
| Number of top preferences among Selah interventions, n (%) | | | | | .429 | .985 |
| No preference | 21.8% (17) | 18.0% (18) | 15.2% (17) | 11.3% (12) | | |
| 1 top preference | 74.4% (58) | 74.0% (74) | 75.9% (85) | 82.1% (87) | | |
| 2 tied preferences | 3.8% (3) | 8.0% (8) | 8.9% (10) | 6.6% (7) | | |
| Body Mass Index, mean (SD) | 31.2 (7.3) | 30.0 (6.6) | 31.1 (7.3) | 30.3 (7.2) | .615 | .975 |
| Self-endorsed high cholesterol, n (%) | | | | | .036 | .405 |
| Never or missing | 73.1% (57) | 52.0% (52) | 60.7% (68) | 56.6% (60) | | |
| Yes, current or history | 26.9% (21) | 48.0% (48) | 39.3% (44) | 43.4% (46) | | |
| Overall life stress at registration, scale range 0-4, mean (SD) | 2.49 (0.75) | 2.31 (0.84) | 2.46 (0.88) | 2.43 (0.88) | .474 | .897 |
| C-SOSI stress symptoms, scale range 0-4, mean (SD) | 1.15 (0.57) | 0.88 (0.55) | 1.03 (0.59) | 0.88 (0.52) | .003 | .156 |
| Self-endorsed single-item anxiety, n (%) | | | | | .016 | .374 |
| Never or missing | 62.8% (49) | 78.0% (78) | 59.8% (67) | 73.6% (78) | | |
| Yes, current or history | 37.2% (29) | 22.0% (22) | 40.2% (45) | 26.4% (28) | | |
| GAD-7 anxiety symptoms, scale range 0-21, mean (SD) | 5.2 (4.0) | 4.1 (4.5) | 5.1 (4.8) | 4.2 (3.9) | .173 | .651 |
| Self-endorsed single item depression, n (%) | | | | | .001 | .101 |
| Never or missing | 60.3% (47) | 76.0% (76) | 50.9% (57) | 68.9% (73) | | |
| Yes, current or history | 39.7% (31) | 24.0% (24) | 49.1% (55) | 31.1% (33) | | |

**Supplemental Table B8.** Baseline characteristic balance between treatment conditions before and after propensity score adjustment for HRV outcomes in pooled trial and observational participants

| | Unadjusted Mean (SD) or % (n) | | | | *P-value* | |
|---|---|---|---|---|---|---|
| | Stress Proofing | Daily Examen | Mindfulness-Based Stress Reduction | Waitlist | Before adjustment | After adjustment |
| Age (in years), mean (SD) | 51.2 (10.9) | 53.5 (12.3) | 52.9 (10.4) | 52.9 (10.1) | .784 | .920 |
| Sex, n (%) | | | | | .108 | .256 |
| Male | 31.1% (14) | 55.6% (25) | 44.6% (29) | 51.0% (26) | | |
| Female | 68.9% (31) | 44.4% (20) | 55.4% (36) | 49.0% (25) | | |
| Race & ethnicity, n (%) | | | | | .878 | .895 |
| Single-racial, non-Latinx white | 93.3% (42) | 91.1% (41) | 93.8% (61) | 90.2% (46) | | |
| All other ethnicities, including bi-/multi-racial | 6.7% (3) | 8.9% (4) | 6.2% (4) | 9.8% (5) | | |
| Marital & habitation status, n (%) | | | | | .741 | .786 |
| Not married, or married but separated/divorcing | 15.6% (7) | 8.9% (4) | 12.3% (8) | 15.7% (8) | | |
| Married or cohabitating with partner | 84.4% (38) | 91.1% (41) | 87.7% (57) | 84.3% (43) | | |
| Any children living at home, n (%) | | | | | .722 | .914 |
| No | 53.3% (24) | 51.1% (23) | 43.1% (28) | 47.1% (24) | | |
| Yes | 46.7% (21) | 48.9% (22) | 56.9% (37) | 52.9% (27) | | |
| Full-time clergy in UMC, n (%) | | | | | .761 | .990 |
| No | 15.6% (7) | 17.8% (8) | 10.8% (7) | 13.7% (7) | | |
| Yes | 84.4% (38) | 82.2% (37) | 89.2% (58) | 86.3% (44) | | |
| Number of congregations appointed to, n (%) | | | | | .577 | .571 |
| Not appointed to a local congregation | 24.4% (11) | 11.1% (5) | 15.4% (10) | 17.6% (9) | | |
| 1 congregation | 53.3% (24) | 73.3% (33) | 67.7% (44) | 66.7% (34) | | |
| 2+ congregations | 22.2% (10) | 15.6% (7) | 16.9% (11) | 15.7% (8) | | |
| Number of congregants pastored, n (%) | | | | | .116 | .438 |
| Not appointed to a local congregation | 31.1% (14) | 13.3% (6) | 18.5% (12) | 19.6% (10) | | |
| 1-149 people in worship per week | 51.1% (23) | 68.9% (31) | 58.5% (38) | 45.1% (23) | | |
| 150+ people in worship per week | 17.8% (8) | 17.8% (8) | 23.1% (15) | 35.3% (18) | | |
| Hours per week worked as clergy, mean (SD) | 47.3 (12.6) | 44.6 (12.3) | 46.7 (12.7) | 49.2 (13.9) | .392 | .923 |

| | | | | | | |
|---|---|---|---|---|---|---|
| Alcoholic drink intake, scale range 0-5, mean (SD) | 1.42 (1.25) | 1.40 (1.44) | 1.32 (1.32) | 1.31 (1.32) | .969 | .956 |
| Caffeinated beverage intake, scale range 0-4, mean (SD) | 1.78 (0.88) | 1.73 (0.96) | 1.57 (0.87) | 1.65 (0.91) | .635 | .663 |
| Metabolic equivalents (METs) per week, mean (SD) | 75.1 (63.0) | 72.7 (100.6) | 39.1 (61.6) | 64.6 (66.9) | .036 | .670 |
| Practicing Daily Examen 3+ times a week at registration, n (%) | | | | | | |
| No | 97.8% (44) | 95.6% (43) | 98.5% (64) | 96.1% (49) | | |
| Yes | 2.2% (1) | 4.4% (2) | 1.5% (1) | 3.9% (2) | | |
| Practicing Mindfulness 3+ times a week at registration, n (%) | | | | | .102 | .403 |
| No or missing | 97.8% (44) | 86.7% (39) | 96.9% (63) | 96.1% (49) | | |
| Yes | 2.2% (1) | 13.3% (6) | 3.1% (2) | 3.9% (2) | | |
| Number of top preferences among Selah interventions, n (%) | | | | | .912 | .990 |
| No preference | 20.0% (9) | 20.0% (9) | 20.0% (13) | 17.6% (9) | | |
| 1 top preference | 77.8% (35) | 73.3% (33) | 75.4% (49) | 80.4% (41) | | |
| 2 tied preferences | 2.2% (1) | 6.7% (3) | 4.6% (3) | 2.0% (1) | .912 | .990 |
| Body Mass Index, mean (SD) | 30.4 (7.1) | 29.6 (6.3) | 31.1 (7.8) | 30.3 (6.7) | .774 | .936 |
| Self-endorsed high cholesterol, n (%) | | | | | | |
| Never or missing | 75.6% (34) | 60.0% (27) | 63.1% (41) | 58.8% (30) | | |
| Yes, current or history | 24.4% (11) | 40.0% (18) | 36.9% (24) | 41.2% (21) | | |
| Overall life stress at registration, scale range 0-4, mean (SD) | 2.56 (0.78) | 2.58 (0.89) | 2.45 (0.92) | 2.57 (1.04) | .852 | .953 |
| C-SOSI stress symptoms, scale range 0-4, mean (SD) | 1.11 (0.56) | 0.98 (0.62) | 0.96 (0.53) | 0.86 (0.58) | .173 | .477 |
| Self-endorsed single-item anxiety, n (%) | | | | | .359 | .846 |
| Never or missing | 64.4% (29) | 73.3% (33) | 58.5% (38) | 70.6% (36) | | |
| Yes, current or history | 35.6% (16) | 26.7% (12) | 41.5% (27) | 29.4% (15) | | |
| GAD-7 anxiety symptoms, scale range 0-21, mean (SD) | 5.0 (3.7) | 4.7 (5.0) | 4.5 (4.3) | 4.2 (4.1) | .821 | .894 |
| Self-endorsed single-item depression, n (%) | | | | | .163 | .982 |
| Never or missing | 62.2% (28) | 68.9% (31) | 49.2% (32) | 64.7% (33) | | |
| Yes, current or history | 37.8% (17) | 31.1% (14) | 50.8% (33) | 35.3% (18) | | |

**Supplemental Table B9.** Between arm, mixed effects regression estimated differences in outcomes between immediate intervention and waitlist control by follow-up time point for trial period participants, using estimates from multiply imputed data (Sensitivity Analysis)

| | Time Point | Stress Proofing | Daily Examen | Mindfulness-based Stress Reduction |
|---|---|---|---|---|
| **Survey outcomes** | | | | |
| C-SOSI (scale range 0-4) | 12 weeks | -0.26 [-0.38, -0.13] | -0.08 [-0.22, 0.05] | -0.29 [-0.42, -0.16] |
| | 24 weeks | -0.22 [-0.42, -0.02] | -0.20 [-0.36, -0.03] | -0.34 [-0.49, -0.20] |
| GAD-7 (scale range 0-21) | 12 weeks | -1.27 [-2.30, -0.24] | -0.49 [-1.47, 0.50] | -1.71 [-2.65, -0.78] |
| | 24 weeks | -1.11 [-2.71, 0.50] | -1.02 [-2.24, 0.20] | -1.76 [-2.85, -0.66] |
| PHQ-8 (scale range 0-24) | 12 weeks | -1.83 [-3.33, -0.33] | -1.25 [-2.60, 0.10] | -1.95 [-3.15, -0.75] |
| | 24 weeks | -0.98 [-2.97, 1.02] | -1.18 [-2.67, 0.32] | -2.06 [-3.51, -0.61] |
| **HRV outcomes** | | | | |
| MESOR (unit = 1 millisecond) | 12 weeks | 0.52 [-3.39, 4.43] | 1.62 [-2.44, 5.68] | 3.80 [0.50, 7.10] |
| Amplitude (unit = 1 millisecond) | 12 weeks | 1.33 [-1.41, 4.06] | 1.34 [-0.78, 3.47] | 2.20 [-0.17, 4.56] |

Abbrev: C-SOSI = Calgary Symptoms of Stress Inventory; GAD-7 = Generalized Anxiety Disorder-7; PHQ-8 = Patient Health Questionnaire-8; HRV = Heart rate variability; MESOR = Midline estimating statistic of rhythm

**Supplemental Table B10.** Between arm, mixed effects regression estimated differences in outcomes between immediate intervention and waitlist control by follow-up time point for trial period participants, using pooled trial period and observational data (Sensitivity Analysis)

| | Sample size | | | Stress Proofing | | Daily Examen | | Mindfulness-Based Stress Reduction | |
|---|---|---|---|---|---|---|---|---|---|
| | Participants | Observations | Time Point | Unadjusted | Adjusted | Unadjusted | Adjusted | Unadjusted | Adjusted |
| **Survey outcomes** | | | | | | | | | |
| C-SOSI (scale range 0-4) | 276 | 780 | 12 weeks | -0.24 [-0.34, -0.15] | -0.26 [-0.37, -0.16] | -0.06 [-0.19, 0.07] | -0.07 [-0.20, 0.06] | -0.25 [-0.35, -0.16] | -0.27 [-0.36, -0.18] |
| | | | 24 weeks | -0.25 [-0.41, -0.10] | -0.35 [-0.49, -0.21] | -0.15 [-0.32, 0.02] | -0.21 [-0.36, -0.06] | -0.30 [-0.42, -0.17] | -0.36 [-0.49, -0.24] |
| GAD-7 (scale range 0-21) | 276 | 780 | 12 weeks | -1.20 [-2.04, -0.36] | -1.44 [-2.34, -0.55] | -0.55 [-1.44, 0.35] | -0.60 [-1.49, 0.28] | -1.64 [-2.40, -0.88] | -1.76 [-2.53, -0.99] |
| | | | 24 weeks | -1.13 [-2.44, 0.18] | -1.85 [-3.03, -0.66] | -0.87 [-2.07, 0.32] | -1.20 [-2.31, -0.09] | -1.78 [-2.81, -0.74] | -2.18 [-3.24, -1.11] |
| PHQ-8 (scale range 0-24) | 272 | 660 | 12 weeks | -1.81 [-2.91, -0.70] | -2.01 [-3.17, -0.85] | -1.19 [-2.40, 0.02] | -1.17 [-2.41, 0.06] | -1.99 [-3.08, -0.91] | -2.11 [-3.20, -1.02] |
| | | | 24 weeks | -1.56 [-2.65, -0.46] | -1.95 [-3.12, -0.78] | -1.25 [-2.45, -0.04] | -1.37 [-2.68, -0.06] | -1.65 [-2.77, -0.53] | -1.88 [-3.06, -0.70] |
| **HRV outcomes** | | | | | | | | | |
| MESOR (unit = 1 millisecond) | 170 | 324 | 12 weeks | -1.91 [-4.99, 1.17] | -0.53 [-3.58, 2.53] | 1.03 [-3.07, 5.14] | 1.31 [-2.54, 5.15] | 2.85 [-0.26, 5.95] | 3.40 [0.43, 6.36] |
| Amplitude (unit = 1 millisecond) | 170 | 324 | 12 weeks | 0.76 [-1.31, 2.83] | 1.05 [-1.09, 3.19] | 1.48 [-0.40, 3.37] | 1.48 [-0.27, 3.23] | 2.14 [0.32, 3.96] | 2.24 [0.32, 4.16] |

Abbrev: C-SOSI = Calgary Symptoms of Stress Inventory; GAD-7 = Generalized Anxiety Disorder-7; PHQ-8 = Patient Health Questionnaire-8; HRV = Heart rate variability; MESOR = Midline estimating statistic of rhythm

**Supplemental Table B11.** Baseline characteristics by unique preferences vs. no unique preference for trial participants and pooled trial/observational participants

| | **Trial** | | | **Pooled Trial and Observational** | | |
|---|---|---|---|---|---|---|
| | **No unique preference** | **Had (and received) unique preference** | ***p*-value** | **No unique preference** | **Had (and received) unique preference** | ***p*-*value*** |
| | (N = 81) | (N = 174) | | (N = 122) | (N = 232) | |
| **Age (in years)** | | | 0.907 | | | 0.723 |
| Mean (SD) | 53.8 (11.6) | 54.0 (11.0) | | 53.8 (11.2) | 53.3 (11.6) | |
| **Sex, n (%)** | | | 0.490 | | | 0.502 |
| Female | 41 (50.6%) | 80 (46.0%) | | 64 (52.5%) | 113 (48.7%) | |
| Male | 40 (49.4%) | 94 (54.0%) | | 58 (47.5%) | 119 (51.3%) | |
| **Race/Ethnicity, n (%)** | | | 0.010 | | | 0.012 |
| White and not Latinx | 69 (85.2%) | 162 (93.1%) | | 101 (82.8%) | 214 (92.2%) | |
| African American and not Latinx | 5 (6.2%) | 10 (5.7%) | | 10 (8.2%) | 12 (5.2%) | |
| Asian-American/Pacific Islander, Native American, Latinx, bi/multi-racial, and other | 7 (8.6%) | 2 (1.1%) | | 11 (9.0%) | 6 (2.6%) | |
| **Marital & habitation status, n (%)** | | | 0.534 | | | 0.132 |
| Not married, separated, or divorced | 10 (12.3%) | 17 (9.8%) | | 20 (16.4%) | 25 (10.8%) | |
| Married or cohabitating | 71 (87.7%) | 157 (90.2%) | | 102 (83.6%) | 207 (89.2%) | |
| **Any children living at home, n (%)** | | | 0.499 | | | 0.335 |
| No | 39 (50.0%) | 95 (54.6%) | | 61 (52.1%) | 133 (57.6%) | |
| Yes | 39 (50.0%) | 79 (45.4%) | | 56 (47.9%) | 98 (42.4%) | |
| **Clergy appointment, n (%)** | | | 0.917 | | | 0.979 |
| Pastoral charge | 67 (82.7%) | 143 (82.2%) | | 99 (81.1%) | 188 (81.0%) | |
| Extension or other | 14 (17.3%) | 31 (17.8%) | | 23 (18.9%) | 44 (19.0%) | |
| **Bi-vocational, n (%)** | | | 0.918 | | | 0.633 |
| No | 78 (96.3%) | 168 (96.6%) | | 115 (95.0%) | 223 (96.1%) | |
| Yes | 3 (3.7%) | 6 (3.4%) | | 6 (5.0%) | 9 (3.9%) | |
| **Hours per week worked as full-time clergy** | | | 0.596 | | | 0.549 |
| Mean (SD) | 48.9 (9.9) | 49.7 (10.8) | | 48.6 (10.6) | 49.4 (10.2) | |

| | | | | | | |
|---|---|---|---|---|---|---|
| **Stress from congregation(s)/work[1], [0-3]** | | | 0.291 | | | 0.159 |
| Mean (SD) | 1.8 (0.7) | 1.9 (0.7) | | 1.8 (0.7) | 1.9 (0.7) | |
| **Financial stress, n (%)** | | | 0.271 | | | 0.764 |
| Not at all or slightly stressful | 56 (73.7%) | 116 (66.7%) | | 78 (67.2%) | 159 (68.8%) | |
| Moderately, very, or extremely | 20 (26.3%) | 58 (33.3%) | | 38 (32.8%) | 72 (31.2%) | |
| **Alcoholic drink intake, n (%)** | | | 0.790 | | | 0.961 |
| None | 27 (36.5%) | 59 (33.9%) | | 38 (33.3%) | 75 (32.5%) | |
| Occasional drink (not every week) | 16 (21.6%) | 44 (25.3%) | | 32 (28.1%) | 61 (26.4%) | |
| 1-2 drinks | 10 (13.5%) | 33 (19.0%) | | 16 (14.0%) | 40 (17.3%) | |
| 3-6 drinks | 13 (17.6%) | 23 (13.2%) | | 15 (13.2%) | 33 (14.3%) | |
| about a drink a day | 6 (8.1%) | 10 (5.7%) | | 10 (8.8%) | 16 (6.9%) | |
| more than a drink a day | 2 (2.7%) | 5 (2.9%) | | 3 (2.6%) | 6 (2.6%) | |
| **Self-reported current heavy alcohol use, n (%)** | | | 0.385 | | | 0.082 |
| No | 72 (97.3%) | 169 (98.8%) | | 110 (96.5%) | 225 (99.1%) | |
| Yes | 2 (2.7%) | 2 (1.2%) | | 4 (3.5%) | 2 (0.9%) | |
| **Caffeinated beverage intake per day, n (%)** | | | 0.404 | | | 0.507 |
| None | 6 (8.1%) | 24 (13.8%) | | 10 (8.8%) | 29 (12.6%) | |
| 1 cup | 18 (24.3%) | 41 (23.6%) | | 29 (25.4%) | 56 (24.2%) | |
| 2-3 cups | 36 (48.6%) | 82 (47.1%) | | 53 (46.5%) | 113 (48.9%) | |
| 4-5 cups | 13 (17.6%) | 20 (11.5%) | | 19 (16.7%) | 25 (10.8%) | |
| 6 or more cups | 1 (1.4%) | 7 (4.0%) | | 3 (2.6%) | 8 (3.5%) | |
| **Metabolic equivalents (METs) per week** | | | 0.015 | | | 0.001 |
| Mean (SD) | 48.1 (71.8) | 75.5 (87.8) | | 43.4 (67.4) | 74.0 (87.5) | |
| **Body Mass Index (BMI)** | | | 0.001 | | | 0.010 |
| Mean (SD) | 32.8 (8.3) | 29.8 (6.1) | | 32.0 (7.8) | 30.0 (6.3) | |
| **Obesity, n (%)** | | | 0.020 | | | 0.117 |
| Not obese (BMI <30) | 33 (40.7%) | 98 (56.3%) | | 54 (44.3%) | 123 (53.0%) | |
| Obese (BMI 30+) | 48 (59.3%) | 76 (43.7%) | | 68 (55.7%) | 109 (47.0%) | |
| **Hypertension, n (%)** | | | 0.263 | | | 0.133 |
| No (including missing) | 57 (70.4%) | 110 (63.2%) | | 88 (72.1%) | 149 (64.2%) | |
| Yes, current or history | 24 (29.6%) | 64 (36.8%) | | 34 (27.9%) | 83 (35.8%) | |
| **Diabetes, n (%)** | | | 0.119 | | | 0.056 |

| | | | | | | |
|---|---|---|---|---|---|---|
| No (including missing) | 67 (82.7%) | 156 (89.7%) | | 102 (83.6%) | 210 (90.5%) | |
| Yes, current or history | 14 (17.3%) | 18 (10.3%) | | 20 (16.4%) | 22 (9.5%) | |
| **PHQ-8 depression symptoms sum score, [0-24]** | | | 0.991 | | | 0.555 |
| Mean (SD) | 5.4 (4.8) | 5.4 (4.5) | | 5.6 (5.0) | 5.3 (4.5) | |
| **Depression screens, n (%)** | | | 0.783 | | | 0.542 |
| Negative (PHQ-8 <10) | 68 (84.0%) | 142 (82.6%) | | 97 (79.5%) | 189 (82.2%) | |
| Positive (PHQ-8 10+) | 13 (16.0%) | 30 (17.4%) | | 25 (20.5%) | 41 (17.8%) | |
| **C-SOSI stress symptoms mean score, [0-4]** | | | 0.672 | | | 0.813 |
| Mean (SD) | 1.0 (0.6) | 1.0 (0.5) | | 1.0 (0.6) | 1.0 (0.5) | |
| **GAD-7 anxiety sum score, [0-21]** | | | 0.682 | | | 0.795 |
| Mean (SD) | 4.6 (4.8) | 4.8 (4.3) | | 4.8 (4.8) | 4.6 (4.2) | |
| **HRV MESOR** | | | 0.929 | | | 0.998 |
| Mean (SD) | 24.6 (13.1) | 24.4 (17.3) | | 25.1 (14.0) | 25.1 (17.5) | |

[1] From November 2019 to registration for Trial participants; from February 2020 to registration for Observational participants

**Supplemental Figure B5.** Subgroup analysis of heterogeneity of treatment effects on depression by preference type

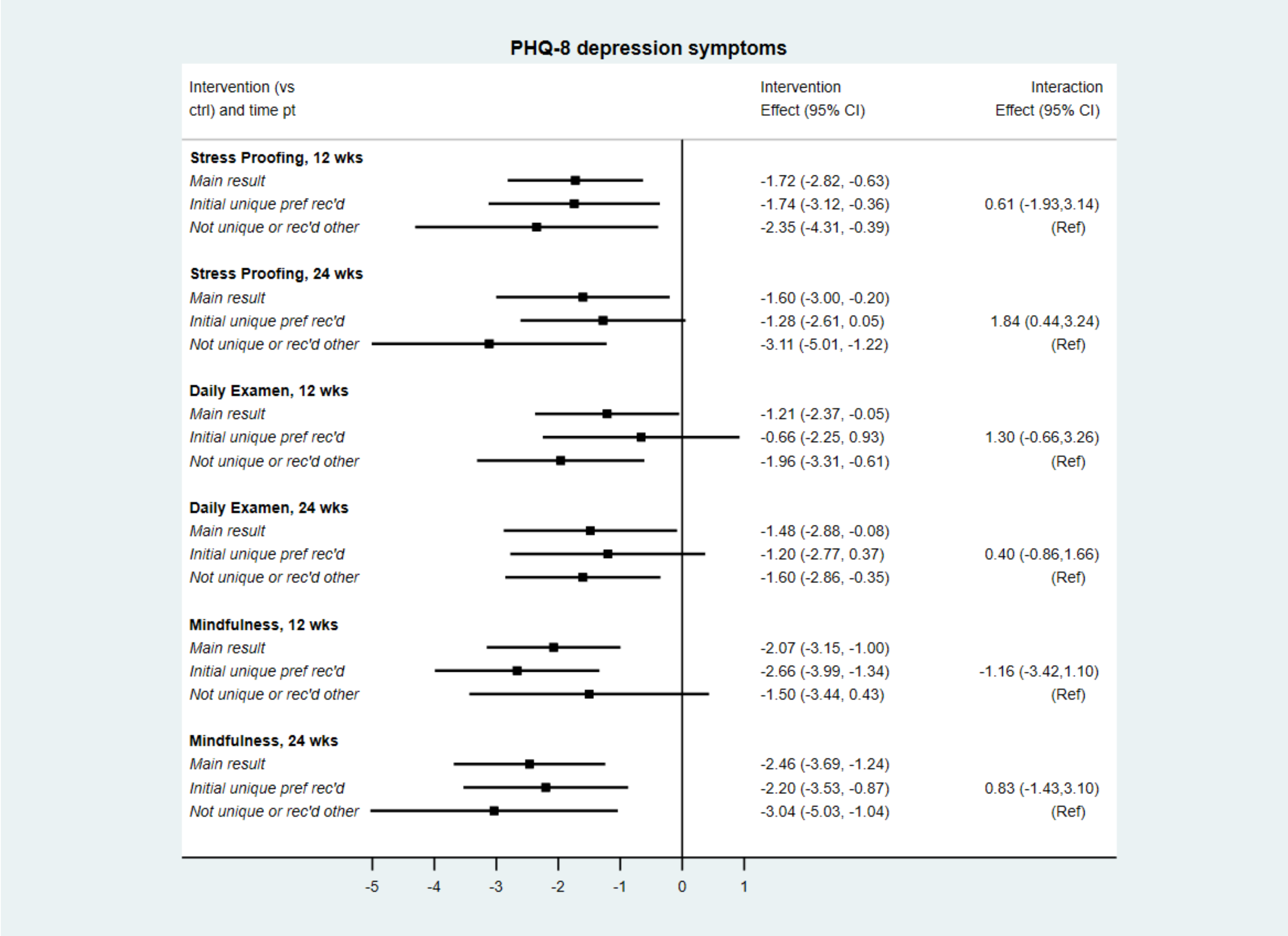

**Supplemental Table B12.** Intra-class correlation coefficients for clustering due to group delivery of intervention estimated using mixed effects regression for trial period participants

| | ***Unadjusted*** | ***Adjusted*** |
|---|---|---|
| ***Survey outcomes*** | | |
| **C-SOSI** | 0.061 | 0.000 |
| **GAD-7** | 0.023 | 0.000 |
| **PHQ-8** | 0.035 | 0.007 |
| ***HRV outcomes*** | | |
| **MESOR** | 0.000 | 0.000 |
| **Amplitude** | 0.000 | 0.000 |

#### CONSORT-SPI 2018 Checklist

| SECTION | ITEM # | CONSORT-SPI 2010 | CONSORT-SPI 2018 | REPORTED ON PAGE # |
|---|---|---|---|---|
| **TITLE AND ABSTRACT** | | | | |
| | 1a | Identification as a randomised trial in the title§ | | 1 |
| | 1b | Structured summary of trial design, methods, results, and conclusions (for specific guidance see CONSORT for Abstracts)§ | Refer to CONSORT extension for social and psychological intervention trial abstracts | 3 |
| **INTRODUCTION** | | | | |
| Background and Objectives | 2a | Scientific background and explanation of rationale § | | 4-6 |
| | 2b | Specific objectives or hypotheses § | If pre-specified, how the intervention was hypothesied to work | 7 |
| **METHODS** | | | | |
| Trial Design | 3a | Describe of trial design (such as parallel, factorial), including allocation ratio § | If the unit of random assignment is not the individual, please refer to CONSORT for Cluster Randomized Trials | 7 |
| | 3b | Important changes to methods after trial commencement (such as eligibility criteria), with reasons | | 9 |
| Participants | 4a | Eligibility criteria for participants§ | When applicable, eligibility criteria for settings and those delivering the interventions | 8 |
| | 4b | Settings and locations where the data were collected | | 10 |
| Interventions | 5 | The interventions for each group with sufficient details to allow replication, including how and when they are actually administered § | | 10-12 |
| | 5a | | Extent to which interventions were actually delivered by providers and taken up by participants as planned | 10-12 |
| | 5b | | Where other informational materials about delivering the intervention can be accessed | 10 |

| | | | | |
|---|---|---|---|---|
| | 5c | | When applicable, how intervention providers were assigned to each group | N/A |
| Outcomes | 6a | Completely defined pre-specified outcomes, including how and when they were assessed§ | | 13-14 |
| | 6b | Any changes to trial outcomes after the trial commenced, with reasons | | N/A |
| Sample Size | 7a | How sample size was determined§ | | 16 |
| | 7b | When applicable, explanation of any interim analyses and stopping guidelines | | N/A |
| **RANDOMISATION** | | | | |
| Sequence generation | 8a | Method used to generate the random allocation sequence | | 9 |
| | 8b | Type of randomisation; detail of any restriction (such as blocking and block size)§ | | N/A |
| Allocation concealment mechanism | 9 | Mechanism used to implement the random allocation sequence, describing any steps taken to conceal the sequence until interventions were assigned§ | | 9 |
| Implementation | 10 | Who generated the random allocation sequence, who enrolled participants, and who assigned participants to interventions§ | | 9-10 |
| Awareness of assignment | 11a | Who was aware of intervention assignment after allocation (for example, participants, providers, those assessing outcomes), and how any masking was done | | 10 |
| | 11b | If relevant, description of the similarity of interventions | | N/A |
| Analytical methods | 12a | Statistical methods used to compare group outcomes§ | How missing data were handled, with details of any imputation method | 17, 19 |

| | | | | |
|---|---|---|---|---|
| | 12b | Methods for additional analyses, such as subgroup analyses, adjusted analyses, and process evaluations | | 18 |
| **RESULTS** | | | | |
| Participant flow (a diagram is strongly recommended) | 13a | For each group, the numbers randomly assigned, receiving the intended intervention, and analysed for the outcomes§ | Where possible, the number approached, screened, and eligible prior to random assignment, with reasons for non-enrolment | Supp Fig B1 |
| | 13b | For each group, losses and exclusions after randomisation, together with reasons§ | | Supp Fig B1 |
| Recruitment | 14a | Dates defining the periods of recruitment and follow-up | | 10 |
| | 14b | Why the trial ended or was stopped | | N/A |
| Baseline data | 15 | A table showing baseline characteristics for each group§ | Include socioeconomic variables where applicable | Table 1 |
| Numbers analysed | 16 | For each group, number included in each analysis and whether the analysis was by original assigned groups§ | | Tables 1-2; Supplemental Figures B1, B2, and B10; and Supplemental Tables B2 and B3 |
| Outcomes and estimation | 17a | For each outcome, results for each group, and the estimated effect size and its precision (such as 95% confidence interval)§ | Indicate availability of trial data | Table 2, page 15 2.6.4 for data availability |
| | 17b | For binary outcomes, the presentation of both absolute and relative effect sizes is recommended | | N/A |
| Ancillary analyses | 18 | Results of any other analyses performed, including subgroup analyses, adjusted analyses, and process evaluations, distinguishing pre-specified from exploratory | | Page 11; Table 2; Figure 1 |
| Harms | 19 | All important harms or unintended effects in each group (for specific guidance see CONSORT for Harms) | | N/A |
| **DISCUSSION** | | | | |
| Limitations | 20 | Summarize the main results (including an overview of concepts, themes, and types of evidence available), link to the | Trial limitations, addressing sources of potential bias, imprecision, and, if relevant, multiplicity of analyses | 24-27; 29-30 |

| | | | | |
|---|---|---|---|---|
| | | review questions and objectives, and consider the relevance to key groups. | | |
| Generalisability | 21 | Discuss the limitations of the scoping review process. | Generalisability (external validity, applicability) of the trial findings§ | 29 |
| Interpretation | 22 | Provide a general interpretation of the results with respect to the review questions and objectives, as well as potential implications and/or next steps. | Interpretation consistent with results, balancing benefits and harms, and considering other relevant evidence | 31 |
| **IMPORTANT INFORMATION** | | | | |
| Registration | 23 | Registration number and name of trial registry | | Abstract, 8 |
| Protocol | 24 | Where the full trial protocol can be accessed, if available | | 8, 10 |
| Declaration of Interests | 25 | Sources of funding and other support; role of funders | Declaration of any other potential interests | Title page |
| Stakeholder investments | 26a | | Any involvement of the intervention developer in the design, conduct, analysis, or reporting of the trial | 12 |
| | 26b | | Other stakeholder involvement in trial design, conduct, or analyses | N/A |
| | 26c | | Incentives offered as part of the trial | 15 |

This table lists items from the CONSORT 2010 checklist (with some modifications for social and psychological intervention trials) and additional items in the CONSORT-SPI 2018 extension. Empty rows in the 'CONSORT-SPI 2018' column indicate that there is no extension to the CONSORT 2010 item

*We strongly recommended that the CONSORT-SPI 2018 Explanation and Elaboration (E&E) document be reviewed when using the CONSORT-SPI 2018 checklist for important clarifications on each item

§An extension item for cluster trials exists for this CONSORT 2010 item

**Citations**

Montgomery, P., Grant, S., Mayo-Wilson, E., Macdonald, G., Michie, S., Hopewell, S., & Moher, D. (2018). Reporting randomised trials of social and psychological interventions: the CONSORT-SPI 2018 Extension. *Trials*, *19*(1), 407.

Grant, S., Mayo-Wilson, E., Montgomery, P., Macdonald, G., Michie, S., Hopewell, S., & Moher, D. (2018). CONSORT-SPI 2018 Explanation and Elaboration: guidance for reporting social and psychological intervention trials. *Trials*, *19*(1), 406.
